# Predicting cognitive function three months after surgery in patients with a glioma

**DOI:** 10.1101/2024.10.08.24315076

**Authors:** Sander Martijn Boelders, Bruno Nicenboim, Elke Butterbrod, Wouter de Baene, Eric Postma, Geert-Jan Rutten, Lee-Ling Ong, Karin Gehring

**Author notes:** **Corresponding author:** Karin Gehring, PhD, Department of Neurosurgery, Elisabeth-Tweesteden Hospital/Cognitive Neuropsychology, Tilburg University, P.O. Box 90153, Warandelaan 2, Tilburg, The Netherlands, 5000 LE,.

## Abstract

**Introduction:** Patients with a glioma often suffer from cognitive impairments both before and after anti-tumor treatment. Ideally, clinicians can rely on predictions of post-operative cognitive functioning for individual patients based on information obtainable before surgery. Such predictions would facilitate selecting the optimal treatment considering patients’ onco-functional balance.

**Method:** Cognitive functioning three months after surgery was predicted for 317 patients with a glioma across eight cognitive tests. Nine multivariate Bayesian regression models were used following a machine-learning approach while employing pre-operative neuropsychological test scores and a comprehensive set of clinical predictors obtainable before surgery. Model performances were compared using the Expected Log Pointwise Predictive Density (ELPD), and pointwise predictions were assessed using the Coefficient of Determination (R²) and Mean Absolute Error. Models were compared against models employing only pre-operative cognitive functioning and the best-performing model was interpreted. Moreover, an example prediction including uncertainty for clinical use was provided.

**Results:** The best-performing model obtained a median R² of 34.20%. Individual predictions, however, were uncertain. Pre-operative cognitive functioning was the most influential predictor. Models including clinical predictors performed similarly to those using only pre-operative functioning (ΔELPD 14.4±10.0, ΔR² −0.53%.).

**Conclusion:** Post-operative cognitive functioning cannot yet reliably be predicted from pre-operative cognitive functioning and the included clinical predictors. Moreover, predictions relied strongly on pre-operative cognitive functioning. Consequently, clinicians should not rely on the included predictors to infer patients’ cognitive functioning after treatment. Moreover, it stresses the need to collect larger cross-center multimodal datasets to obtain more certain predictions for individual patients.

**Importance of the study:** Patients with a glioma often suffer from cognitive impairments both before and after anti-tumor treatment. Ideally, clinicians would be able to rely on predictions of cognitive functioning after treatment for individual patients based on information that is obtainable before surgery. Such predictions would facilitate selecting the optimal treatment considering patients’ onco-functional balance and could improve patient counseling. First, our study shows that cognitive functioning three months after surgery cannot be reliably predicted from pre-operative cognitive functioning and the included clinical predictors, with pre-operative cognitive functioning being the most important predictor. Consequently, clinicians should not rely on the included predictors to infer individual patients’ cognitive functioning after surgery. Second, results demonstrate how individual predictions resulting from Bayesian models, including their uncertainty estimates, may ultimately be used in clinical practice. Third, our results show the importance of collecting additional predictors and stress the need to collect larger cross-center multimodal datasets.

**Key points:**

- Cognitive functioning after treatment cannot yet reliably be predicted
- Pre-operative cognitive functioning was the most important predictor
- Additional predictors and larger cross-center datasets are needed

## Introduction

Patients with a glioma often suffer from cognitive impairments, both before and after anti-tumor treatment^1,2^, which may contribute to a decreased quality of life^3–5^. Cognitive impairments after anti-tumor treatment are likely caused by the damage inflicted by the tumor before surgery^6,7^, the surgical resection^8^, and adjuvant therapies^9,10^. Moreover, cognitive functioning after anti-tumor treatment has been related to numerous patient characteristics, such as age, education, and medicine use^11^, and cognitive functioning before surgery appears to be one of the strongest indicators of post-operative functioning^12,13^. Unfortunately, the exact mechanisms by which glioma affect cognitive functioning after treatment remain poorly understood.

The consideration of cognitive functioning is becoming increasingly important in determining the optimal treatment in view of patients’ onco-functional balance. This onco-functional balance refers to weighing the oncological benefit of treatment against its adverse side effects on the functional status and quality of life of the patient^14^. Ideally, clinicians would be able to use predictions of cognitive functioning after treatment to facilitate selecting the optimal treatment^13,15–17^.

Unfortunately, achieving accurate predictions of cognitive functioning at the individual level is challenging due to two sources of uncertainty: aleatoric and epistemic uncertainty^18^. Here, aleatoric uncertainty refers to the inherent randomness present in most real-world settings, such as the variability in measurements of cognitive functioning^19^. Epistemic uncertainty stems from an incomplete understanding of the causal mechanisms behind observed data, such as how surgery impacts cognitive functioning. Given that predictions may be unreliable due to aleatoric and epistemic uncertainty, it is essential to use methods to quantify uncertainty in individual predictions such that clinicians know when predictions can be relied upon^20^.

Bayesian models offer two main advantages. First, they can be used to model the uncertainty in individual predictions^21^. Bayesian models do this by learning distributions of possible values for each parameter, rather than point estimates. By combining these parameter distributions with the predictors for a new data point, Bayesian models produce a probability distribution of potential outcomes. This distribution can be used to obtain a point estimate and reflects the uncertainty in the prediction.

Second, Bayesian models allow for incorporating prior knowledge into parameter estimates using priors. These priors represent our beliefs about the parameters before having seen any data, potentially improving model performance^22^. Even though the mechanisms by which glioma affect cognitive functioning are poorly understood, weakly informative priors can still be used to provide some guidance to the models. Bayesian models already have significant traction for making predictions in clinical applications^22^ and for describing neurological functions^19,23^, and are becoming increasingly accessible^24^.

Predictions of cognitive functioning after treatment could aid in selecting the optimal treatment. Unfortunately, previous studies could only partially explain cognitive functioning after treatment at the individual level and are limited by a small sample size, a small number of included predictors, and do not model uncertainty in individual predictions^13^. To address these limitations, the current study aims to predict cognitive functioning after treatment in a large sample of patients with a glioma (n=317) using Bayesian models employing a comprehensive set of predictors available before surgery. The current study is an extension to our previous study where we employed machine-learning models to predict pre-operative cognitive functioning using the same set of predictors^25^.

## Method

### Participants

Patients were included when they had an oligodendroglioma or astrocytoma (WHO grade 2, 3, and 4) and underwent elective surgery between 2010 and 2019 at the Elisabeth-TweeSteden Hospital, Tilburg, The Netherlands, and had a valid pre-operative cognitive screening as performed during clinical care. Patients were not included when they had reduced testability (e.g. no serious visual or motor deficits) for the neuropsychological screening, were under 18, had a progressive neurological disease, or had a psychiatric or acute neurological disorder within the previous two years. This study was part of a protocol registered with the Medical Ethics Committee Brabant (file number NW2020-32). This is the same sample as (in part) included in^12,25–29^.

### Interview and cognitive testing

Informed consent was obtained prior to performing a standardized interview. This interview was performed to collect age, sex, and education (the Dutch Verhage scale), and to measure symptoms of anxiety and depression using the Dutch translation of the Hospital Anxiety and Depression Scale (HADS)^30^ for use as predictors.

Cognitive functioning was assessed immediately before and three months after surgical resection of the tumor using the CNS Vital Signs (CNS VS)^31^ computerized neuropsychological test battery. The psychometric properties of this battery were shown to be comparable to the pen-and-paper tests that it is based on in patients with various neuropsychiatric disorders and healthy individuals^32–35^. A well-trained technician (neuropsychologist or neuropsychologist in training) provided test instructions and reported on the validity of each test. Requirements for the validity included the patient understanding the test, showing sufficient effort, having no vision or motor impairments that significantly affected task performance, and the absence of any (external) distractions. Invalid tests were excluded on a test-by-test basis. Test scores were calculated from the CNS VS results according to the formulas presented in Appendix 1 and were defined such that a higher score represents a better performance.

### Clinical characteristics

Five of the variables used for prediction were collected from patients’ electronic medical records. This set consisted of the involved hemisphere, the use of antiepileptic drugs, comorbidities, the ASA score (assessment of the patient’s physical status before surgery^36^), and the symptoms the patient presented with. These presenting symptoms were categorized into five binary categories indicating whether the symptom was present or not: behavioral/self-reported cognitive problems, language problems, epilepsy/loss of consciousness, motor deficits (including paresis), and headache.

Three tumor characteristics were also included as predictors. These were the tumor grade which describes the malignancy of the tumor (classified according to the WHO guidelines as used at the time of treatment^37,38^), histopathological diagnosis (oligodendroglioma, astrocytoma as based on cell origin/molecular markers), and IDH1 mutation status. Note that we used the measured values for these tumor characteristics whereas they can only be estimated preoperatively^39^.

In our clinical practice, the IDH mutation status of patients with a grade 4 glioblastoma aged over 55 is not always tested due to the very low incidence rate of IDH mutant gliomas for these patients^40–42^. Therefore, missing IDH mutation statuses for this subset of patients were set to wild-type. A detailed explanation is provided in Appendix 2.

### Tumor volume and location

Tumor volume and location were also used as predictors. Tumors were segmented automatically from routine MRI scans. All segmentations were manually validated and redone semi-automatically when deemed incorrect. For low-grade gliomas, the tumor region was defined as the hyperintense area on the FLAIR scan, while for high-grade gliomas, it was defined as the hyperintense area on the T1 contrast scan. Additional details regarding segmentation are provided in Appendix 3. Tumor volume was quantified by the number of voxels (mm³) in the segmentation. Tumor location was determined by calculating the percentage of overlap between segmentations and the four lobes individually for each hemisphere^26^.

### Analysis

#### Follow-up Participation

Not all included patients with a valid pre-operative screening returned for or were able to complete the follow-up neuropsychological assessment three months after surgery. Therefore, these patients were not included in the prediction models. The number of patients without a valid follow-up measurement, along with their reasons was reported. Additionally, patient characteristics and cognitive test scores were statistically compared between those with and without a valid follow-up measurement. This was done using either a t-test, Mann–Whitney U test, or Chi-Square test. No corrections for multiple testing were applied given the descriptive nature of these analyses.

#### Modeling

##### Variable reduction

The complete set of predictors is listed in Table 1. Although the models used in this study can handle large numbers of predictors due to the use of shrinkage priors (see model specification below), an excessive number of predictors can hinder model convergence and increase uncertainty in individual predictions. Therefore, the number of predictors was reduced by considering the number of patients in different categories (at least 10% per category), variance inflation factors (< 0.5), pairwise correlations (> 0.6 for to-be-combined variables), and the interpretability of combined predictors^25^.

**Table 1:**
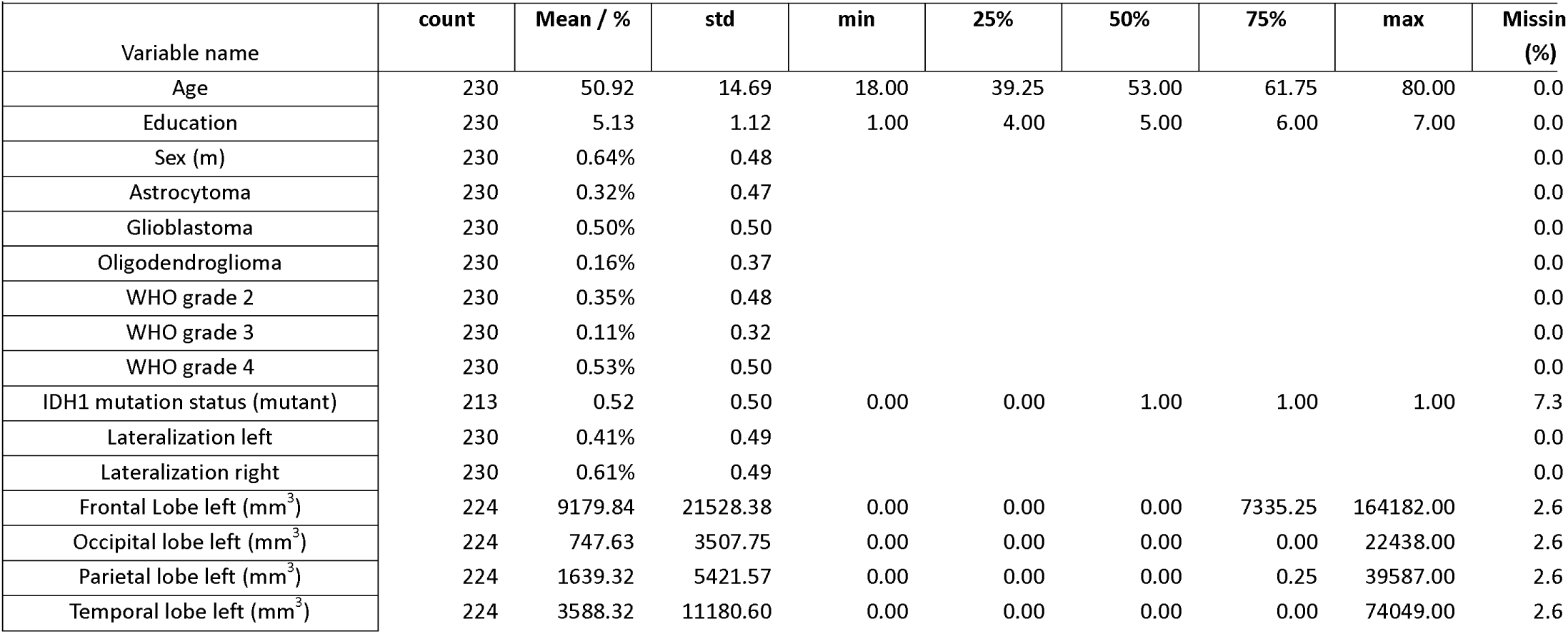
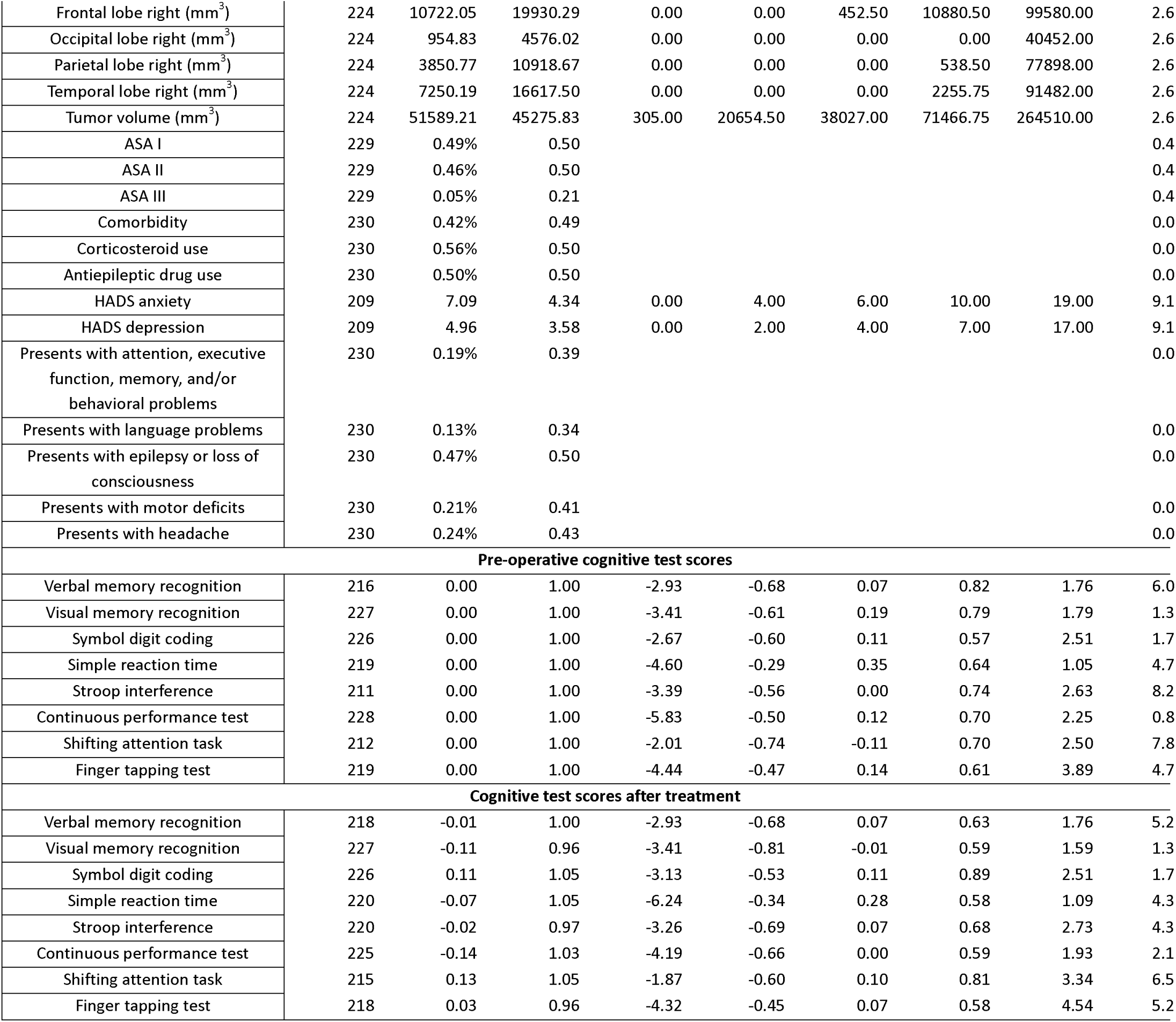
Sample characteristics of the sample used for model fitting. Pre-operative test scores were normalized to have zero mean and unit variance. Cognitive test scores three months after surgery were scaled relative to the pre-operative test scores.

##### Preprocessing

All predictors were normalized to have a mean of zero and a standard deviation of one to ensure all predictors contribute equally during model fitting and to aid the interpretation of model parameters. Test scores three months after surgery were normalized relative to pre-operative scores to ensure they were on the same scale, facilitating interpretation. Moreover, pre-operative scores of patients without valid follow-up screenings, who were not included in the models, were normalized relative to those with valid follow-ups for descriptive purposes. As no statistical analyses are performed in the current study, cognitive test scores were not normalized relative to healthy participants, corrected for effects of age, sex, and education as found in healthy participants, nor corrected for test-retest effects.

##### Model specification

Three Bayesian models (models 1, 2, and 3) were evaluated for predicting cognitive functioning three months after surgery. These models were the following:

**Model 1** was a multiple multivariate linear regression model (i.e., a model with multiple predictors and multiple outcomes). A multivariate approach was used to allow for the joint estimation of model parameters for the eight different test scores.

**Model 2** was similar to the first but included interaction effects between predictors and the histopathological diagnoses (oligodendroglioma, astrocytoma, or glioblastoma). These interactions were included as predictors of cognitive function, while related, may vary across different diagnoses^43^.

**Model 3** was also a multiple multivariate linear model but allowed coefficients to differ between histopathological diagnoses using partial pooling. This method allows coefficients to vary across groups while pulling them toward the population average.

All three models were evaluated while modeling residual correlations between the test scores as cognitive test scores are known to be correlated^44^. Moreover, all models were fitted with **(a)** no (additional) interaction effects, **(b)** an (additional) interaction effect between age and tumor volume, or **(c)** an (additional) interaction effect between education level and tumor volume. These interaction effects were added since evidence for the role of tumor volume by itself on postoperative cognitive function is mixed^11^ and may be moderated by proxies of neuroplasticity and cognitive reserve such as age and education level^45^.

Models were defined using the Bayesian Regression Models using Stan (BRMS) package (v2.20.1)^24,46,47^. A formal description of the models including the BRMS syntax is presented in Appendix 4. Models were fitted using the Hamiltonian Monte Carlo algorithm in STAN (v2.21)^48^. Missing test scores were estimated within the Bayesian models themselves. Missing predictors were imputed before fitting the models with multiple imputation using MICE^49^ (v3.16.0). Thirty different imputed datasets were created and models were fitted individually on each of the imputed datasets. Afterward, the model parameters were pooled to account for the uncertainty in imputation.

For all models, weakly informative priors were used for the coefficients, intercept, residuals, and random effects instead of informative priors for two reasons. First, previous studies employed various neuropsychological tests, which differ in both their sensitivity and the cognitive domains they measure. Consequently, model parameters may not translate to the tests used in this study. Second, we do not expect our model parameters to be independent, complicating the determination of informative priors. Alongside the weakly informative priors, expectations regarding the number of non-zero coefficients and the magnitude of coefficients were set using horseshoe priors^50,51^ because of the small sample-to-variable ratio. For a detailed rationale behind each individual prior used we refer to Appendix 5. To verify that the priors correctly modeled our expectations, prior predictive checks were performed for each model and described for the best-performing model (see below).

#### The role of pre-operative cognitive functioning

To assess the added value of clinical predictors beyond pre-operative cognitive functioning, three additional models were fitted using only pre-operative cognitive functioning as predictors. These models were labeled as 1d, 2d, and 3d and mirror the structure of models 1a, 2a, and 3a respectively while not including the clinical predictors. Note that models 2b and 3b still include the interaction effects with or structure across different histopathological diagnoses.

#### Model convergence and evaluation

To explore how well the sampling process explored the parameter space, the effect size (ESS) was evaluated. To determine if the model converged, the Rhat values were inspected. An ESS of above 1000 and a Rhat below 1.05 were interpreted as sufficient.

Models were compared using the expected log pointwise predictive density as determined using the expected leave-one-out cross-validation (ELPD-LOO)^52^ with Pareto smoothed importance sampling (PSIS)^53^. The ELPD-LOO is a Bayesian measure for model comparison that approximates the out-of-sample generalizability of model predictions based on the full posterior distributions. The best-performing model was defined as having the highest ELPD-LOO.

To facilitate comparison with studies using frequentist machine-learning models, point-wise predictions resulting from the best-performing model were evaluated using 10-fold cross-validation. Here, point-wise predictions were defined as the mean of the posterior predictive distribution, and were evaluated using the frequentist versions of the mean absolute error (MAE) and coefficient of determination (R^2^) score. Normalization and imputation of the predictors were performed within the cross-validation loop to prevent information leakage^54^. To assess the added value of clinical predictors, point-wise predictions were additionally evaluated for the model that obtained the highest ELPD-LOO while only employing pre-operative cognitive functioning (and potentially tumor histopathology), and the plain multivariate model that only employed pre-operative cognitive functioning (model 1d). To evaluate whether the best-performing model is a good fit for the observed data, the posterior predictive distributions resulting from this model were visualized.

#### Sensitivity to selected priors

To ensure the priors were only weakly informative, fitted model parameters including their credibility intervals (i.e. the posterior distributions) were inspected for the best-performing model. Moreover, to test the sensitivity to the selected priors and to distinguish between the effect of the horseshoe prior and all other priors, model performance was compared against three additional versions of the best-performing model. These were the same model with only the default priors in BRMS, and versions with weakly informative priors where either the horseshoe prior or all weakly informative priors were replaced with their default.

#### Model interpretation and application

To interpret the relationships captured by the best-performing model, its fitted model parameters including their credibility intervals were inspected. To interpret the certainty of the individual out-of-sample predictions, the amount of uncertainty in predictions as obtained using the 10-fold cross-validation (i.e. posterior predictive distributions) was described. Last, to inspect whether the model made any systematic errors, the point-wise out-of-sample predictions were plotted against the measured values.

To illustrate the application of Bayesian models and their uncertainty estimates can be applied in clinical practice, an out-of-sample prediction resulting from the best-performing model was visualized. This was achieved by showing the point estimate, the posterior predictive distribution describing the uncertainty, and the true measured value. The prediction was selected to have a standard deviation in the posterior predictive distribution closest to the population median, thus having a median amount of uncertainty. Multiple example predictions for all outcome measures selected to differ in their amount of uncertainty were provided as an appendix.

The Bayesian Analysis Reporting Checklist by Kruschke^55^ was followed and is provided as an online supplement. Moreover, documented R (v4.0.4)^56^ code and dummy data are provided as an online supplement.

## Results

### Descriptive statistics and follow-up participation

A total of 317 patients were included in the study. Eighty of these patients did not participate in the three-month follow-up. The reasons for not participating in the follow-up were: not responding to, not showing up for, or canceling the appointment without reporting a reason (n=24); being clinically unable to show up for or perform the assessment (23); having passed away (12); being treated in a different hospital (7); logistical reasons (2); or undergoing a re-resection (1). For eleven patients, the reason could not be determined retrospectively. Finally, for seven of the remaining 237 follow-up measurements, all tests in the battery were deemed invalid by the test technician. Descriptive statistics for the remaining sample (n=230) are provided in Table 1. Moreover, descriptive statistics for the sample that did not participate in the follow-up or had a follow-up measurement that was not deemed valid are presented in Appendix 6.

Patients who did not participate in the follow-up or had an invalid follow-up measurement more often had a comorbidity (chi=6.89, p=0.009), and had lower ASA scores (U=1166, p=0.021). Note that no differences in age (U=11191, p=0.103), sex (chi=0.018, p= 0.893), and education (U=10267, p=0.708) were found. Regarding cognitive test scores, patients who did not participate in the follow-up or had an invalid follow-up measurement had a significantly lower score pre-operatively on the measure of verbal memory recognition (U=7315, p=0.020), the symbol digit coding task (T=-4.50, p=0.000), the measure of Simple reaction time (U=6906, p=0.000), and the shifting attention task (U=6693, p=0.013), but not the other four measures (all p’s>0.127).

### Variable reduction

Based on the variance inflation factor, pairwise correlations, and number of patients per category, tumor lateralization was grouped into right-lateralized and left-lateralized + bilateral; tumor grades were grouped into low-grade (grade 2) and high-grade (grade 3 + 4); ASA scores were grouped into ASA I and ASA II + III; use of antiepileptic drugs was merged with ‘presenting with epilepsy or loss of consciousness’; and HADS anxiety and depression were combined. The resulting set of predictors used as predictors is the same as used in our previous study^25^.

### Model convergence and evaluation

The prior predictive check for the best-performing model (model 2c, see below) is reported in Appendix 7 and was highly similar for all other models. The prior predictive check showed that the simulated data covered a wide but reasonable range of outcomes. This indicates that the selected priors were weakly informative.

BRMS did not report problems with model fitting. The Rhat values generally were below 1.05 with a small number of exceptions ranging up to 1.13, indicating good convergence. The ESS for the bulk and tail of the distributions for the different models generally were above 1000 with a small number of exceptions as low as 575 and 836 for the bulk and tail ESS respectively, indicating effective sampling.

Table 2 (part 1) presents the ELPD-LOO for each model including the difference relative to the best-performing model. Model 2c achieved the best performance (ELPD-LOO = −1624) and includes an interaction effect between education and tumor volume and interactions with the histopathological diagnosis. The five runner-ups (model 2a, 2b, 3a, 3b, and 3c) performed similarly with decreases in ELPD-LOO ranging from −2.2 to −11.0. These differences were smaller than the standard deviations in this difference which were between 7.9 and 14.7. Therefore, we cannot distinguish between these models according to their ELPD-LOO. Models that performed partial pooling (Models 3) performed the worst, with a decrease in ELPD-LOO of at least −127.6 (SE=17.7). Finally, there was no clear effect of including the interaction effect between tumor volume and age or education level. This can be seen from the small differences between variants a, b, and c of the different models.

**Table 2:**
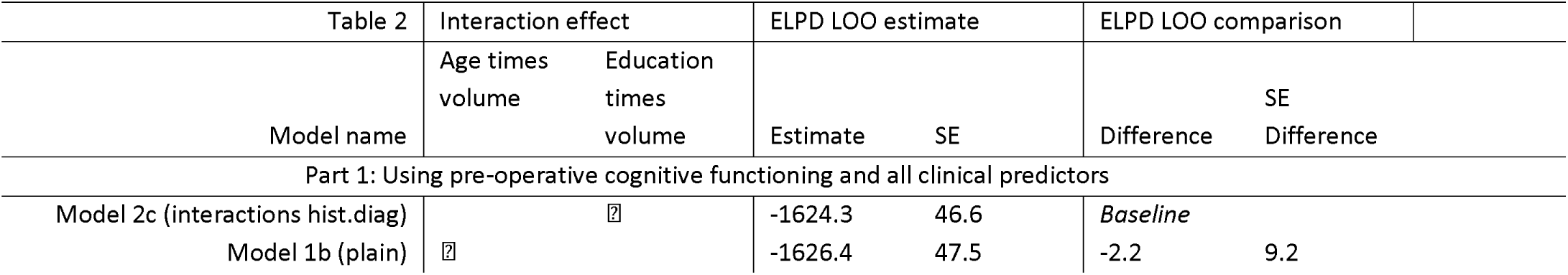
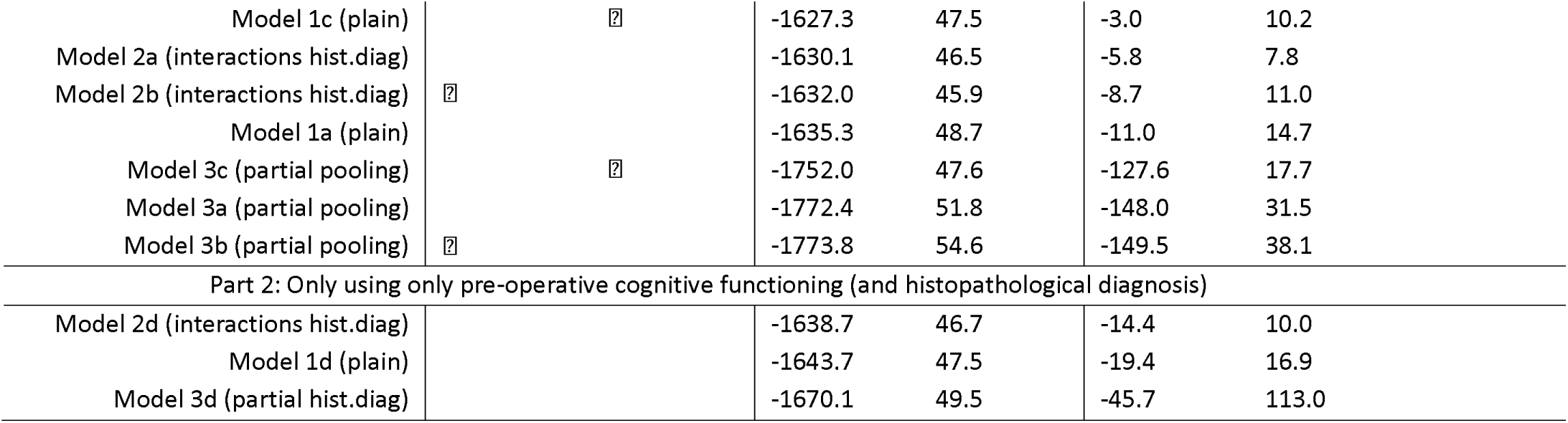
Model performance sorted from best to worst individually for models including the clinical predictors (part 1), and models not including clinical predictors (part 2). Performance is described as the Expected Log Predictive Density - Leave-One-Out (ELPD-LOO) and the standard error of this estimate is reported. Moreover, the difference of all models relative to the best-performing model is reported including the expected standard error of this difference. hist.diag: Histopathological diagnosis

Table 2 (part 2) presents the performance of models 1d, 2d, and 3d which only employed pre-operative cognitive functioning (and histopathological diagnosis) and were evaluated to test the added value of the clinical predictors. Of these models, model 2d performed best with an ELPD-LOO of −1638.7. Comparing its performance to the best-performing model overall (2c), which has the same structure, only a slight decrease in performance is observed with a difference of −14.4 (SE=10.0).

Table 3 describes the out-of-sample prediction performance of the pointwise predictions using the mean absolute error (MAE) and the coefficient of determination (R^2^). These results show that the best-performing model as determined using the ELPD-LOO (model 2c) obtained a median R^2^ of 34.20% of variance and a median MAE of 0.599. Performance for the individual tests ranged between an R^2^ of 13.77% and an MAE of 0.693 for the Stroop interference ratio and an R^2^ of 73.22% and an MAE of 0.420 for the symbol digit coding task.

**Table 3:** The mean absolute error (MAE) and the coefficient of determination (R2) individually for each test score and the median across the different test scores. Hist.diag: Histopathological diagnosis.

|  | Model 2c (Best model):<br>interactions hist.diag and<br>education times size |  | Model 2d: Best model when<br>using only pre-operative<br>functioning and<br>histopathology: |  | Model 1d: Model using only<br>pre-operative functioning: |  |
| --- | --- | --- | --- | --- | --- | --- |
| Table 3 | $R^2$ score | MAE | $R^2$ score | MAE | $R^2$ score | MAE |
| Verbal memory recognition | 26.78% | 0.661 | 28.26% | 0.660 | **28.73%** | **0.659** |
| Visual memory recognition | **27.93%** | **0.666** | 27.71% | 0.674 | 28.78% | 0.670 |
| Symbol digit coding | **73.22%** | **0.420** | 69.47% | 0.441 | 69.47% | 0.441 |
| Simple reaction time | 25.48% | 0.602 | **28.27%** | **0.590** | 27.32% | 0.592 |
| Stroop interference ratio | 13.77% | **0.693** | 12.59% | 0.706 | **13.96%** | 0.697 |
| Continuous performance test | **51.06%** | 0.539 | 49.91% | **0.537** | 49.49% | 0.543 |
| Shifting attention task | 48.64% | 0.597 | **48.97%** | **0.587** | 48.89% | 0.589 |
| Finger tapping test | 40.47% | 0.508 | **41.19%** | **0.499** | 41.01% | 0.503 |
| Median | 34.20% | 0.599 | 34.73% | **0.589** | **34.89%** | 0.590 |

The median R^2^ score of the best-performing model (2c) and its MAE were lower when compared to both models 1d and 2d which only relied on pre-operative cognitive functioning (and histopathological diagnosis). This difference, however, is very small with a change of 0.54 percentage points in R2 and a change of 0.01 in MAE.

When considering individual test measures, the best-performing model (Model 2c) obtained the highest R^2^ score and MAE for the measure of verbal memory recognition and the symbol digit coding task. Moreover, this model obtained the highest R^2^ score for the continuous performance test and the highest MAE for the Stroop interference ratio.

To evaluate whether the best-performing model (2c) accurately describes the distribution of the observed data, the posterior predictive checks for this model are visualized in Appendix 8. This figure shows that the simulated data matches the observed data for most draws from the model parameters and training data. This indicates that the model was able to adequately describe the observed data. For most tests, however, and especially the measure of simple reaction time, the model was not able to completely capture the skewness.

### Sensitivity to selected priors

The parameter estimates after model fitting (i.e. the posterior distributions) of the best-performing model (model 2c) are visualized in Appendix 9. This visualization shows that they were within the specified priors, indicating that the priors were suitable.

Two of the three sensitivity checks as performed for model 2c did not converge. These were the variant with all default priors, and the variant with weakly-informative priors but no horseshoe prior. The variant with all default priors except for the horseshoe prior converged and performed slightly worse when compared to model 2c with a difference in ELDP-LOO of −12.53 (SE=9.85). This shows that the horseshoe prior was crucial for model convergence while the weakly-informative priors only had a small positive impact on model fit.

### Model interpretation and application

For the estimated model parameters for the best-performing model (2c), we refer back to Appendix 9. Note that the relationships captured by the model are solely descriptive of how the model obtains its predictions.

Results showed that the most important predictor of a given measure of cognitive functioning after treatment was this same measure before treatment with coefficients ranging between 0.35 [95% CI: 0.20, 0.48] and 0.74 [95% CI: 0.63, 0.84] (Appendix 9A). One notable exception from this was the Stroop interference ratio, whose predictions relied mostly on the pre-operative measure of the shifting attention task with a coefficient of 0.23 [95% CI: 0.20, 0.40], followed by the pre-operative measure of the Stroop interference ratio with a coefficient of 0.11 [95% CI: 0.00, 0.26].

The contribution of most clinical predictors was negligible with coefficients generally being around zero with only 3.34% of the coefficients being above |0.05|. Moreover, the credibility intervals associated with these coefficients were large relative to the magnitudes of the coefficients. Additionally, the included interaction effect between education level and size contributed little to the predictions with coefficients of at most |0.02|.

The expected variability in the measures of cognitive functioning after treatment (i.e. standard deviation of the likelihood) ranged between 0.53 [95% CI: 0.0.48, 0.59] for symbol digit coding and 0.84 [95% CI: 0.76, 0.93] for verbal memory recognition (Appendix 9C). The median amount of uncertainty in the resulting predictions as obtained using 10-fold cross-validation (i.e. standard deviations of the posterior predictive distributions) ranged between 0.56 for the symbol digit coding task and 0.94 for the Stroop interference ratio (Appendi× 10).

To inspect whether the model made any systematic errors, out-of-sample predictions were plotted against the measured values in Figure 1. This figure shows that there are no systematic deviations in model performance as can be seen from most points being clustered around the line representing perfect predictions. For Simple reaction time, however, there is some heteroscedasticity. This can be seen from predictions for patients who scored poorly on this measure showing more variance in prediction error.

**Figure 1:**
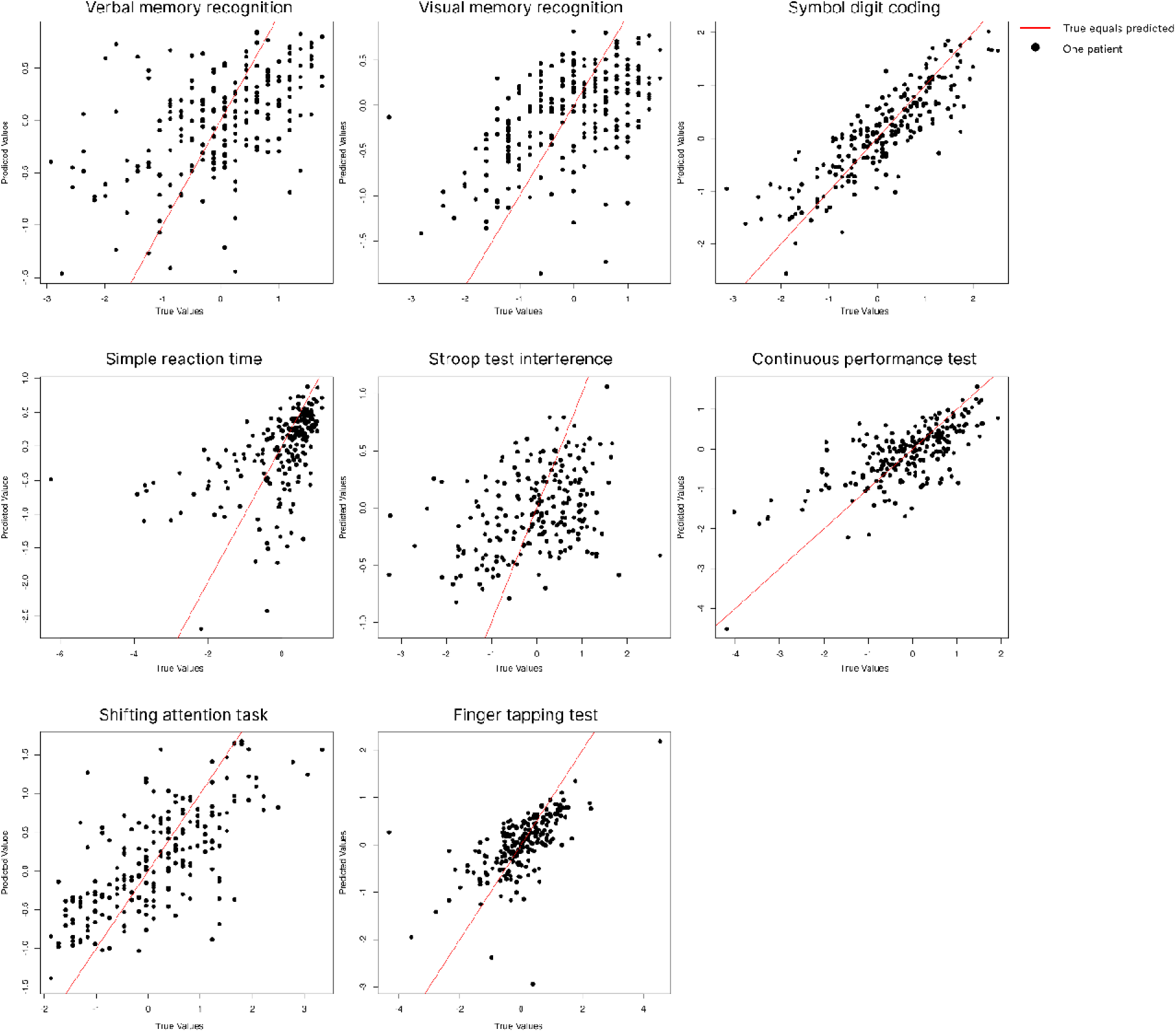
Scatter plots of predicted values obtained using 10-fold cross-validation from the best-performing model (2c) versus the measured values, individually for each outcome measure. Each dot represents a patient, its position along the x-axis represents their measured value, and the position along the y-axis the predicted test score. The red line (x=y) represents perfect predictions, and the distance along the x-axis represents the error of the prediction.

A demonstration of how uncertainty estimates can be used in clinical practice is provided in Figure 2. The large range of outcomes covered by the uncertainty estimate shown in this figure (in blue) indicates that clinicians should not rely on the point estimate (in red) as there is a large chance that it will not be close to the true value (in green). Therefore, clinicians should not rely on this prediction for decision-making and can inform patients we don’t know how the treatment will affect their cognitive functioning. Additional examples for all outcome measures are presented in Appendix 11, showing similar uncertainty in the predictions.

**Figure 2:**
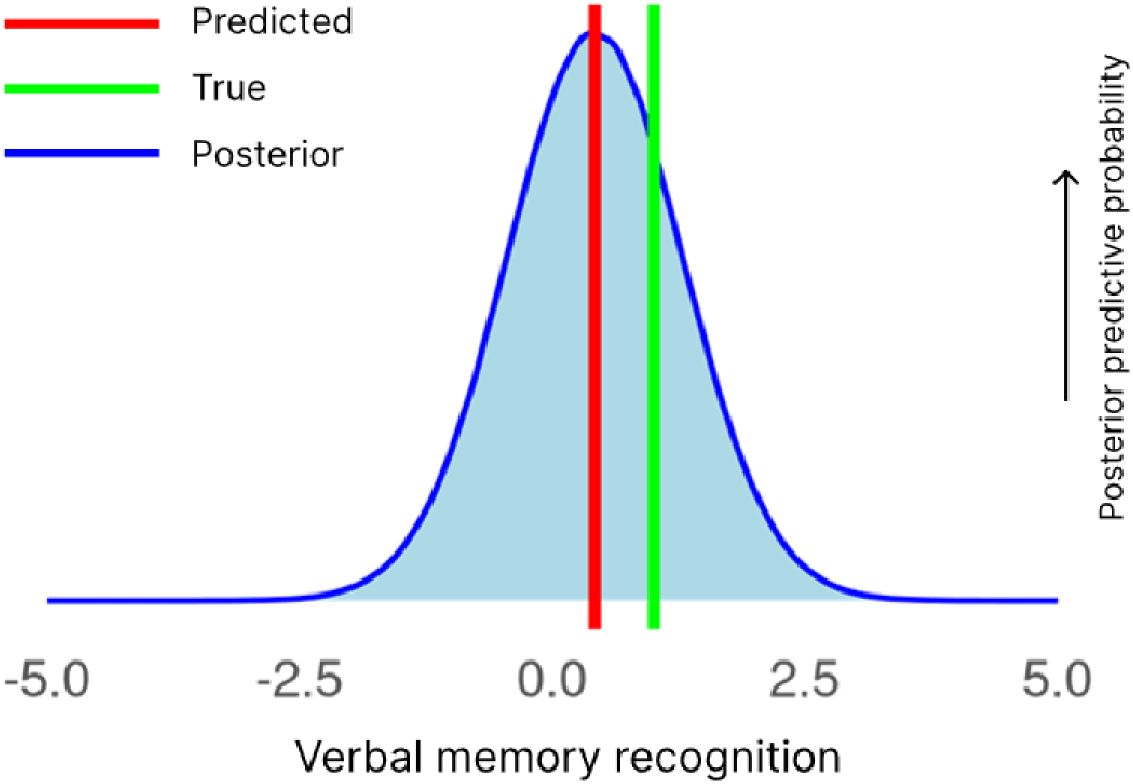
Example predictions of verbal memory recognition obtained using 10-fold cross-validation for the best-performing model (2c). The example prediction was selected to be at the median in terms of the amount of uncertainty in the prediction. The blue distribution represents the posterior predictive distribution resulting from the model, which represents the probability of each outcome, the red line represents the point estimate obtained from this distribution, and the green line represents the measured value. Three example predictions for all cognitive tests can be found in Appendix 11.

## Discussion

Results show that predicting cognitive functioning three months after surgery using pre-operative cognitive functioning and the included clinical predictors is not yet possible. The amount of variance explained in cognitive functioning three months after surgery ranged between 13.77% (Stroop interference ratio) and 73.22% (Symbol digit coding) with a median of 34.20%. Moreover, the uncertainty in individual predictions ranged between a median standard deviation of 0.72 and 0.94 (relative to the population standard deviations). This performance likely is insufficient for clinical application, though further research is needed to establish thresholds for clinical utility. These findings align with the study by Zangrossi and colleagues which explained between 0.81% and 62.41% (median 39.09%) of variance in cognitive performance one week after awake surgery^13^ while employing age, education, and pre-operative neuropsychological test scores.

The best-performing model relied strongly on pre-operative cognitive functioning to predict cognitive functioning three months after surgery, in line with previous studies^12,13^. Moreover, the model using only pre-operative cognitive functioning and interactions with the histopathological diagnosis as predictors (model 2d) performed similarly to the best-performing model overall (2c). Additionally, the included interaction effects between tumor volume and age or education level had a negligible influence on predictions. These findings show that the added value of the clinical predictors and included interaction effects as used in the current study are limited when predicting cognitive functioning after treatment.

The maximum amount of variance that a perfect model can explain is unknown and limited by the aleatoric uncertainty. One part of this aleatoric uncertainty stems from the test-retest reliability. For CNS VS, the test-retest reliability has only been established for healthy participants^35^ and likely is lower for patients with a brain tumor due to cognitive impairments and medication effects. The epistemic uncertainty likely results from relevant predictors that were not included. This set of predictors likely comprises predictors that only are available after treatment and therefore could not be used, such as surgical complications and the adjuvant treatments received^8,10,57–61^, and promising predictors that can be obtained before surgery but are not (yet) routinely collected in our practice including measures of structural and functional connectivity^6,62^ and information regarding edema^57^. Additionally, including different representations of predictors or interactions thereof may improve model performance. Finally, using models that can capture more complex relationships may reduce uncertainty, although this likely requires larger datasets^25^.

The amount of variance explained in cognitive functioning after treatment differed up to 59.4 percentage points between tests. These substantial differences can likely be attributed to three factors. First, some cognitive domains may be more prone to change after surgery, causing the pre-operative test scores to be less informative. Second, the predictive power of the clinical predictors and pre-operative test scores to describe the change in functioning may differ across test scores. Third, the cognitive tasks used differ in their ability to reliably and repeatably measure the cognitive functions they are intended to measure, in line with differences in their test-retest reliability^35^.

The current models assume that the decision for surgery is already made. Ideally, predictive models would avoid such assumptions, thereby enabling clinicians to compare predictions across various treatment decisions. This, however, requires either data from randomized control trials (RCTs)^63^, simulating an RCT from retrospective data^64,65^, or developing casual models^66,67^. Unfortunately, neither was possible as RCTs are undesirable, simulating an RCT requires all confounders influencing the treatment decision to be available, and causal models require the causal mechanism to be known.

Several limitations of the current study should be noted. First, models are solely based on patients with a valid three-month follow-up. Therefore, predictions are only valid if the patient will be able to undergo the follow-up, which is unknown before surgery. Consequently, the current model needs to be paired with a model that predicts whether a patient will complete the follow-up. Second, we used the histopathological diagnosis, WHO grade, and IDH1 status as determined post-operatively while they can merely be estimated pre-operatively^39^, potentially further limiting the accuracy when applied in clinical practice. Third, the sample was gathered during clinical care and therefore did not include patients with severe impairments or in need of immediate surgical intervention. Finally, cognitive assessment was done using a brief computerized test battery which may be somewhat dependent on processing speed and does not measure language function, memory free recall, or visuoconstructive abilities. However, more comprehensive evaluations are not typically conducted during clinical care.

We believe our results to be highly important as they show that clinicians should not rely on the included clinical predictors to infer cognitive functioning three months after surgery. Additionally, our results demonstrate how estimates of uncertainty ultimately can be used in clinical practice to facilitate trust in predictions. Finally, our results show the importance of collecting larger datasets including additional predictors.

This need for larger datasets is especially important when including high-dimensional and noisy data such as structural and functional connectivity^68^. Moreover, the relatively low signal-to-noise ratio in scores resulting from brief neuropsychological screening^19,35^, and the large individual differences between patients add to this need. Therefore, we hope future work will focus on standardizing data collection to obtain larger cross-center multimodal datasets. Such datasets have the potential to significantly improve the ability to predict cognitive functioning at the individual level while allowing models to generalize across centers. This need is being increasingly emphasized by numerous authors (e.g.^1,6,25,69,70^)

Future studies can use information regarding the planned treatment (both primary and adjuvant) to improve predictions and could utilize virtual models of the brain to model the hypothesized effect of the planned surgery^6,71^. Moreover, future work could predict outcomes that are closer to patients daily functioning.

## Conclusion

Predictions of cognitive functioning after treatment could aid in selecting the optimal treatment. The current study aimed to predict cognitive functioning three months after surgery (and adjuvant treatments) on the individual level using a comprehensive set of clinical predictors available before surgery and pre-operative cognitive functioning while employing Bayesian models. While predictions accounted for substantial variance in cognitive functioning three months after surgery, individual predictions were uncertain and likely of insufficient quality for use in clinical practice. Consequently, clinicians should not rely on the included predictors to infer patients’ cognitive functioning after treatment. Pre-operative cognitive functioning was the most influential predictor and models including clinical predictors and pre-operative functioning performed roughly similarly to models using only pre-operative functioning, showing the limited added value of the clinical predictors and interaction effects as used in the current study. The current study further demonstrated how individual predictions including their uncertainty estimates may ultimately be used in clinical practice, allowing models to say ‘I don’t know’ instead of being confidently wrong. Finally, it stresses the need to collect larger cross-center multimodal datasets including additional predictors. Such datasets have the potential to significantly improve the ability to predict cognitive functioning at the individual level while allowing models to generalize across centers.

## Supporting information

Example data

Reporting checklist

## Funding

ZonMw (10070012010006, 824003007).

## Conflict of interest

None to declare

## Authorship

Experimental design: (SB, KG, EP, GR, LLO, BN), acquisition: (KG, GR, EB), analysis: (SB, LLO, KG, BN), interpretation: (SB, LLO, KG, BN, EP, EB, WDB, GR). All authors have been involved in the writing of the manuscript and approved the final version.

## Acknowledgments

We would like to express our gratitude to Sacha van der Donk for her role as data manager on this project

## Data availability

Data described in this work is not publicly available to protect the privacy of patients. All code used in this study is available as supplementary material.

## Notes

### Competing Interest Statement

The authors have declared no competing interest.

### Funding Statement

This study was funded by ZonMw (10070012010006, 824003007).

### Author Declarations

This study was part of a protocol registered with the Medical Ethics Committee Brabant (file number NW2020-32).

