## Appendix for "Predicting cognitive function three months after surgery in patients with a glioma"

### Appendix 1: Formulas used to obtain the test scores

| Name | Description | Formula |
| --- | --- | --- |
| Verbal memory recognition | Memory recognition for words. Fifteen words are presented one at a time. The subject identified the presented words amongst new words. The immediate condition is at the beginning of the test battery and the delayed condition is at the end. | Number of items correct (immediate and delayed recall) |
| Visual memory recognition | Memory recognition for abstract images. Fifteen images are presented one at a time. Subjects identified the presented images amongst new images. The immediate condition is at the beginning of the test battery and the delayed condition is at the end. | Number of items correct (immediate and delayed recall) |
| Symbol digit coding | Eight symbols are presented on the screen with a corresponding number. Given a row of eight randomly ordered symbols, the participant is asked to provide the matching number for two minutes straight. | Correct responses - incorrect responses |
| Simple reaction time | The subject presses the space bar when a word is presented. | -1 * Average reaction time |
| Stroop test interference | The subject presses the space bar if a word is presented and the color of the word does/or does not match its semantic meaning for the congruent and incongruent trials respectively | -1 * (Average reaction time on correct responses for the incongruent trials - Average reaction time on correct responses for the congruent trials) / Average reaction time on correct responses for the congruent trials |
| Shifting attention task | A red circle and a blue square are presented on the screen. Given a third shape, the participant needs to match the shape either by color or shape. | Correct responses - incorrect responses |
| Continuous performance test | The subject responds to a target letter amongst distractors for 5 minutes straight. | -1 * Average reaction time to the target letter |
| Finger tapping test | The participant presses the spacebar as often as possible within ten seconds. This task is performed three times for each hand. | The average number of presses across left and right trails |

*Caption: Formulas used to obtain the cognitive domains*

### Appendix 2: Imputing IDH mutation status

In the current study, imputation of predictors is performed using MICE (Multiple Imputation by Chained Equations), where each missing variable is estimated based on other variables. For patients aged 55 or higher with a grade IV glioblastoma, missing values on IDH mutation status were imputed based on the literature instead of using multiple imputation. This was done as for this subset of patients, the IDH mutation status is known to be wildtype in 96% of cases^1^. Not imputing these values based on prior knowledge implies that the iterative imputer must learn how to predict IDH status for this subset of patients. The imputer would have to learn how to impute these values from the relatively small number of patients aged 55 or higher with a grade 4 glioblastoma patients for whom IDH status is available. Imputing these values using the iterative imputer, therefore, likely would have resulted in less accurate estimations of IDH status when compared to imputing these values based on the literature. The more accurate the predictions of IDH status are, the more likely it is that we can accurately predict cognitive functioning. Therefore, we opted to impute these values based on clinical knowledge.

Note that the goal of imputation differs between predictive modeling and explanatory modeling. In predictive modeling, imputation is done with the goal of obtaining the highest model performance possible. For explanatory modeling, imputation is done with to goal of being able to use participants with missing values without biasing the results of statistical tests.

### Appendix 3: Segmentation models

All anatomical MRI (T1, T1 contrast, T2, Flair) scans were registered to MNI space using affine transformation. Registration was performed using Regaladin^2^ from the NiftyReg package which has been shown to perform well for patients with a primary brain tumor^3^. Skull stripping was performed using HD-BET which is designed to be robust to a variety of different lesions^4^. Tumor size was defined as the FLAIR-enhancing part for low-grade gliomas and the T1 contrast-enhancing part for high-grade gliomas.

Two different models were used for segmentation and the best segmentation was selected manually for each patient. Models used were nnU-Net^5^ as trained on T1, T1c, T2, and Flair scans from the BraTS dataset or subsets thereof^6,7^ and AGU-Net as available in the Radionics tool using T1c images for high-grade gliomas and FLAIR images for low-grade gliomas^8^. All automatic segmentations were manually validated and incorrect segmentations were redone semi-automatically using the snake tool in ITK-Snap^9^.

### Appendix 4a: Formal model specification

For patients $n\in N,$ we have a response matrix $Y_{post}$ of shape $8 x |N|$ representing the eight test scores as measured post-operatively and a vector $X_{p}$ representing the predictors including the eight test scores as measured pre-operatively. Here, vector $X_{p}$ has shape ($|p|+8) x |N|$ where *p* is the set of predictors used and |*p*|=34 for models without added interaction effects (models 1a, 2a, and 3a) and |*p*|=35 for the models with an interaction effect between age or education level and tumor size. Moreover, we have a contrast coding matrix $X_{g}$ of shape $2 x |N|$ representing the histopathological diagnosis. Here, $X_{g} \cup X_{p}= X$ and $X_{p} \cap X_{g}=\emptyset$. Now, given a vector for the intercepts $\alpha$ of length eight, a fixed effect coefficients matrix $\beta_{p}$ of shape $32 x 8$, a coefficients matrix for the histopathological diagnosis $\beta_{g}$ of shape $2 x 8$, and a full covariance matrix of the residuals *Σ* with shape $8 x 8$, we define the multivariate multiple linear regression model as:

$$Model 1: Y\sim MVN\left( \alpha+X_{g}^{T}\beta_{g}^{T} + X_{p}^{T} \beta_{p}, \Sigma\right)$$

Next, given a coefficient matrix for the interaction between histopathological diagnosis and the other predictors $\beta_{\mathrm{interaction}}$ of shape 68$x 8$, we define the model including interaction effects with histopathological diagnosis as:

$$Model 2: Y \sim MVN\left( \alpha+ X_{g}^{T}\beta_{g} + X_{p}^{T}\beta_{p}+{(X_{g}{\boldsymbol{\odot}X}_{p})}^{T}\beta_{\mathrm{interaction}},\Sigma\right)$$

Finally, given a matrix *u_g_* of shape $32 x 8$ representing the random effects specific to group *g*, a vector $\mu_{\alpha g}$ of length eight representing the random intercepts specific to group *g,* and the diagonal matrices $\Sigma_{\mu}$ for the random effects $\mu_{g},\mu_{\alpha g}$ with shape $33 x 33$ individually for each $y$, we define the partial pooling model as:

$$Model 3: Y\sim MVN\left( \alpha+\mu_{\alpha g} + X_{p}^{T}(\beta+\mu_{g}),\Sigma\right)$$

$$\mathrm{where}{(\mu}_{g},\mu_{\alpha g})\sim N\left( 0,\Sigma_{\mu} \right)$$

All models were implemented using Bayesian Regression Models using Stan (BRMS), and the BRMS model syntax for all models can be found in Appendix 3.

### Appendix 4b: BRMS formulas

Given patients $n\in N,$ we have a response matrix $Y_{post}$ of shape $8 x |N|$ representing the eight test scores as measured post-operatively and a vector $X_{p}$ representing the predictors including the eight test scores as measured pre-operatively. Here, vector $X_{p}$ has shape ($|p|+8) x |N|$ where *p* is the set of predictors used and |*p*|=34 for models without added interaction effects (models 1a, 2a, and 3a) and |*p*|=35 for the models with an interaction effect between age or education level and tumor size. Moreover, we have a contrast coding matrix $X_{g}$ of shape $2 x |N|$ representing the histopathological diagnosis. Here, $X_{g} \cup X_{p}= X$ and $X_{p} \cap X_{g}=\emptyset$. Then we have the following BRMS syntax:

1. A multiple multivariate regression model

$$mvbind(Y) | mi() \sim X+ set\_rescor(TRUE)$$

2. A multilevel multiple multivariate regression model including interaction effects with the histopathological diagnosis of the tumor

$$mvbind(Y) | mi() \sim X_{p}* as.factor(g)+ set\_rescor(TRUE)$$

3. A multilevel multiple multivariate regression model conditioned on the histopathological diagnosis with partial pooling of coefficients and intercepts over the different diagnoses

$$mvbind(Y) | mi() \sim X_{p}+ (1 +X_{p} | g)+ set\_rescor(FALSE)$$

### Appendix 5: Prior specification

The output distribution was modeled as a multivariate normal distribution as the post-operative test scores are largely normally distributed and this allows for modeling residual correlations between outcome measures. The prior for the standard error of individual predictions (the diagonal in $\Sigma$) was modeled as a half-normal distribution with a location of 0 and a scale of 3 ${(\sigma}_{\epsilon}\sim\mathcal{N}_{+}\left( 0, 3 \right)$). A scale of 3 was chosen as it is larger than the post-operative test scores standard deviation which ranges between 0.96 and 1.05, thus ensuring that model certainty comes from the data and not from the prior.

The priors for the intercepts were set to be normally distributed with a mean of 0 and a standard deviation of 2 $\left( \alpha\sim\mathcal{N}\left( 0, 2 \right) \right)$. A mean of zero was chosen as the mean of the post-operative test scores is between -0.14 and 0.13 while all predictors were centered around zero. A standard deviation of 2 was chosen to allow intercepts to become larger or smaller than the most extreme outcomes in post-operative test scores, which were between -6.24 and 4.54.

Regarding the residual correlations, the prior for the covariance matrix $\Sigma$ was set to be the Lewandowski-Kurowicka-Joe distribution with the value for the shape set to 6. This was done as we expect most correlations to be around zero while also allowing for moderate correlations of around 0.5. Moreover, we expected all correlations across tests to be lower than the test-retest reliability of the tests themselves, which is at most 0.88 for healthy participants and is likely lower for patients.

For the partial pooling models (model 3), the prior for the standard deviation between parameter estimates across groups (the diagonal of $\Sigma_{u}$) was set to a half normal with a location of 0 and a scale of 0.5 ${(\sigma}_{u}\sim\mathcal{N}_{+}\left( 0, 0.5 \right)$). This was done as we expected coefficients to be small for most parameters as both the predictors and outcome measures were normalized between 0 and 1, leading to small coefficients and even smaller standard deviations in parameter estimates.

Coefficients for each outcome measure were regularized using horseshoe priors^10,11^. Horseshoe priors restrict the complexity of the potential models by shrinking both the magnitude of coefficients and the number of non-zero coefficients. The horseshoe priors were configured to expect 33% of the coefficients to be non-zero ($\beta_{i}\sim\text{Horseshoe}\left( 1, \text{par}\text{\_}\text{ratio}=0.5 \right)$). This was done for two reasons. First, the best models when predicting pre-operative cognitive functioning using the same set of predictors relied weakly on many predictors without relying strongly on any one specific predictor^12^. Second, the current study includes a large number of predictors (n=34) relative to the number of patients used during model fitting (n=230), which may lead to wide and uninformative predictions or problems with model convergence when not using such a shrinkage prior. All other parameters of the horseshoe priors were left at their default.

### Appendix 6: Descriptive statistics of patients who did not participate in the follow-up or had a follow-up measurement that was not deemed valid

| Variable name | **count** | **Mean / %** | **std** | **min** | **25%** | **50%** | **75%** | **max** | **Missing (%)** |
| --- | --- | --- | --- | --- | --- | --- | --- | --- | --- |
| Age | 87 | 53.91 | 13.27 | 23.00 | 47.00 | 57.00 | 63.00 | 76.00 | 0.00 |
| Education | 87 | 5.15 | 1.23 | 2.00 | 4.50 | 5.00 | 6.00 | 7.00 | 0.00 |
| Sex (m) | 87 | 0.66 | 0.48 | 0.00 | 0.00 | 1.00 | 1.00 | 1.00 | 0.00 |
| Astrocytoma | 87 | 0.26 | 0.44 | 0.00 | 0.00 | 0.00 | 1.00 | 1.00 | 0.00 |
| Glioblastoma | 87 | 0.61 | 0.49 | 0.00 | 0.00 | 1.00 | 1.00 | 1.00 | 0.00 |
| Oligodendroglioma | 87 | 0.09 | 0.29 | 0.00 | 0.00 | 0.00 | 0.00 | 1.00 | 0.00 |
| WHO grade 2 | 87 | 0.01 | 0.11 | 0.00 | 0.00 | 0.00 | 0.00 | 1.00 | 0.00 |
| WHO grade 3 | 87 | 0.01 | 0.11 | 0.00 | 0.00 | 0.00 | 0.00 | 1.00 | 0.00 |
| WHO grade 4 | 87 | 0.06 | 0.23 | 0.00 | 0.00 | 0.00 | 0.00 | 1.00 | 0.00 |
| IDH1 mutation status (mutant) | 69 | 0.49 | 0.50 | 0.00 | 0.00 | 0.00 | 1.00 | 1.00 | 20.69 |
| Lateralization left | 87 | 0.46 | 0.50 | 0.00 | 0.00 | 0.00 | 1.00 | 1.00 | 0.00 |
| Lateralization right | 87 | 0.55 | 0.50 | 0.00 | 0.00 | 1.00 | 1.00 | 1.00 | 0.00 |
| Frontal Lobe left (mm^3^) | 87 | 6550.03 | 18022.68 | 0.00 | 0.00 | 0.00 | 2699.00 | 131285.00 | 0.00 |
| Occipital lobe left (mm^3^) | 87 | 1070.68 | 5035.57 | 0.00 | 0.00 | 0.00 | 0.00 | 33466.00 | 0.00 |
| Parietal lobe left (mm^3^) | 87 | 2227.34 | 6921.95 | 0.00 | 0.00 | 0.00 | 6.50 | 42487.00 | 0.00 |
| Temporal lobe left (mm^3^) | 87 | 4857.57 | 13571.96 | 0.00 | 0.00 | 0.00 | 0.00 | 72405.00 | 0.00 |
| Frontal lobe right (mm^3^) | 87 | 5775.20 | 11765.52 | 0.00 | 0.00 | 51.00 | 5029.00 | 66980.00 | 0.00 |
| Occipital lobe right (mm^3^) | 87 | 708.94 | 2718.80 | 0.00 | 0.00 | 0.00 | 0.00 | 21372.00 | 0.00 |
| Parietal lobe right (mm^3^) | 87 | 7548.37 | 17039.48 | 0.00 | 0.00 | 0.00 | 6168.00 | 77783.00 | 0.00 |
| Temporal lobe right (mm^3^) | 87 | 9350.82 | 18114.78 | 0.00 | 0.00 | 0.00 | 5538.00 | 93323.00 | 0.00 |
| Tumor size (mm^3^) | 87 | 52487.20 | 39828.22 | 2856.00 | 24971.00 | 44980.00 | 70330.00 | 259367.00 | 0.00 |
| ASA I | 87 | 0.40 | 0.49 | 0.00 | 0.00 | 0.00 | 1.00 | 1.00 | 0.00 |
| ASA II | 87 | 0.54 | 0.50 | 0.00 | 0.00 | 1.00 | 1.00 | 1.00 | 0.00 |
| ASA III | 87 | 0.06 | 0.23 | 0.00 | 0.00 | 0.00 | 0.00 | 1.00 | 0.00 |
| Comorbidity | 87 | 0.56 | 0.50 | 0.00 | 0.00 | 1.00 | 1.00 | 1.00 | 0.00 |
| Corticosteroid use | 87 | 0.68 | 0.47 | 0.00 | 0.00 | 1.00 | 1.00 | 1.00 | 0.00 |
| Antiepileptic drug use | 87 | 0.51 | 0.50 | 0.00 | 0.00 | 1.00 | 1.00 | 1.00 | 0.00 |
| HADS anxiety | 85 | 7.32 | 4.69 | 0.00 | 4.00 | 6.00 | 11.00 | 16.00 | 2.30 |
| HADS depression | 85 | 4.81 | 3.42 | 0.00 | 2.00 | 5.00 | 7.00 | 16.00 | 2.30 |
| Presents with attention, executive function, memory, and/or behavioral problems | 87 | 0.30 | 0.46 | 0.00 | 0.00 | 0.00 | 1.00 | 1.00 | 0.00 |
| Presents with language problems | 87 | 0.16 | 0.37 | 0.00 | 0.00 | 0.00 | 0.00 | 1.00 | 0.00 |
| Presents with loss of consciousness | 87 | 0.37 | 0.49 | 0.00 | 0.00 | 0.00 | 1.00 | 1.00 | 0.00 |
| Presents with motor deficits | 87 | 0.31 | 0.47 | 0.00 | 0.00 | 0.00 | 1.00 | 1.00 | 0.00 |
| Presents with headache | 87 | 0.21 | 0.41 | 0.00 | 0.00 | 0.00 | 0.00 | 1.00 | 0.00 |
| **Pre-operative cognitive test scores** | | | | | | | | | |
| Verbal memory recognition | 82 | -0.28 | 0.90 | -2.37 | -0.87 | -0.31 | 0.44 | 1.19 | 5.75 |
| Visual memory recognition | 84 | -0.18 | 1.09 | -3.01 | -0.81 | -0.01 | 0.59 | 1.79 | 3.45 |
| Symbol digit coding | 86 | -0.56 | 0.98 | -2.61 | -1.25 | -0.53 | 0.29 | 1.22 | 1.15 |
| Simple reaction time | 87 | -0.38 | 1.13 | -5.88 | -0.84 | -0.01 | 0.42 | 0.88 | 0.00 |
| Stroop interference | 81 | -0.23 | 1.07 | -2.91 | -0.81 | -0.11 | 0.54 | 2.11 | 6.90 |
| Continuous performance test | 86 | -0.16 | 1.20 | -6.01 | -0.64 | 0.02 | 0.64 | 1.62 | 1.15 |
| Shifting attention task | 78 | -0.36 | 0.93 | -1.87 | -1.03 | -0.32 | 0.21 | 1.51 | 10.34 |
| Finger tapping test | 77 | -0.04 | 0.88 | -3.91 | -0.49 | 0.14 | 0.58 | 1.61 | 11.49 |

*Caption: Descriptive statistics for the sample that did not participate in the follow-up or had a follow-up measurement that was not deemed valid. Pre-operative test scores were scaled relative to the pre-operative test scores of patients that did have a valid follow-up measurement.*

### Appendix 7: Prior predictive check

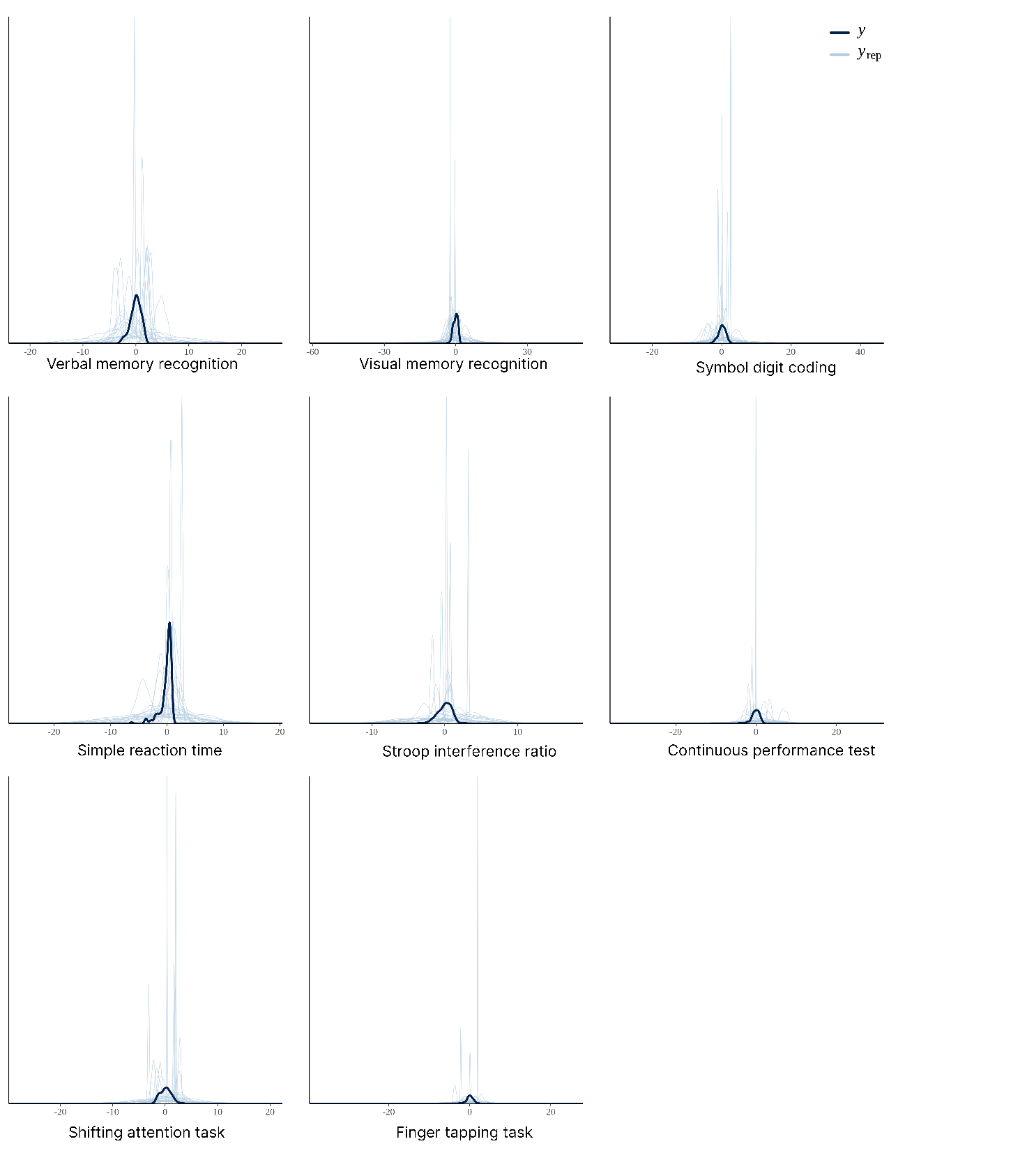

*Caption: Prior predictive check for the best performing model (model 2c), individually for each outcome measure. The light blue lines y_rep_ represents the simulated data and the dark blue line y represents the true output distribution.*

### Appendix 8: Posterior predictive check

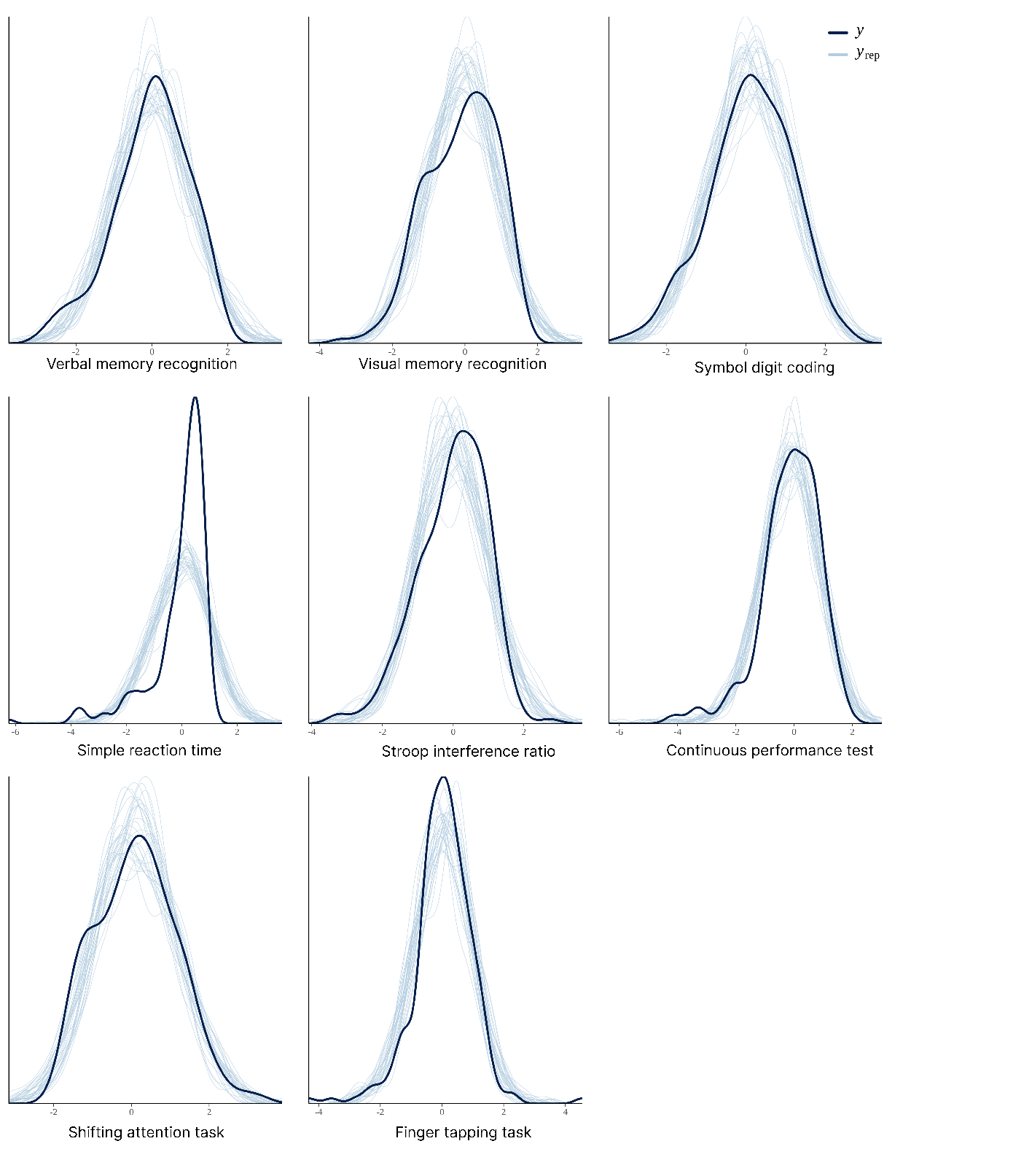

*Caption: Posterior predictive check for the best performing model (model 2c), individually for each outcome measure. The light blue lines y_rep_ represents the simulated data and the dark blue line y represents the true output distribution.*

### Appendix 9: distributions of the parameter estimates (best-performing, Model 2c)

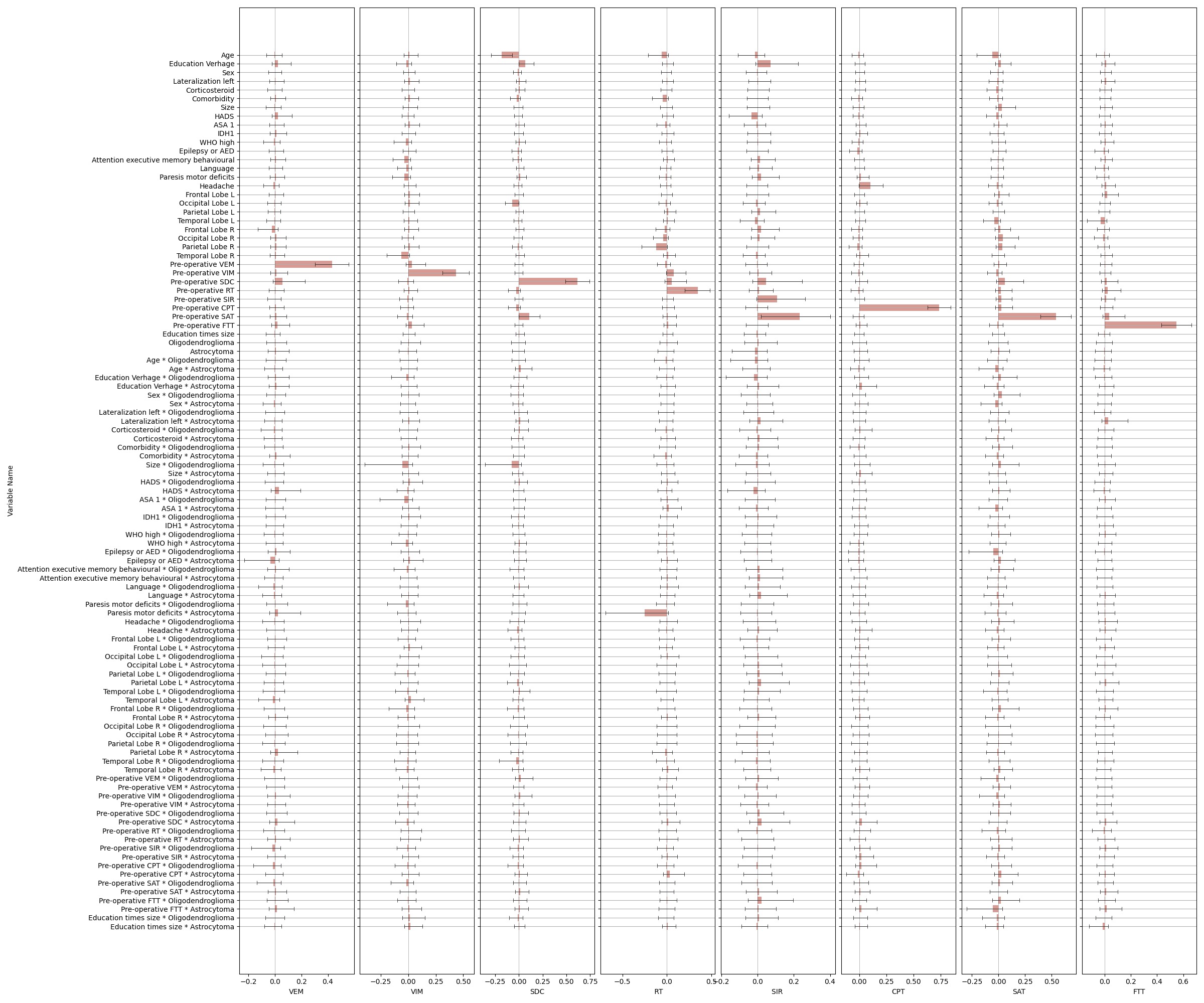

##### A: Coefficients

##### B: Intercepts

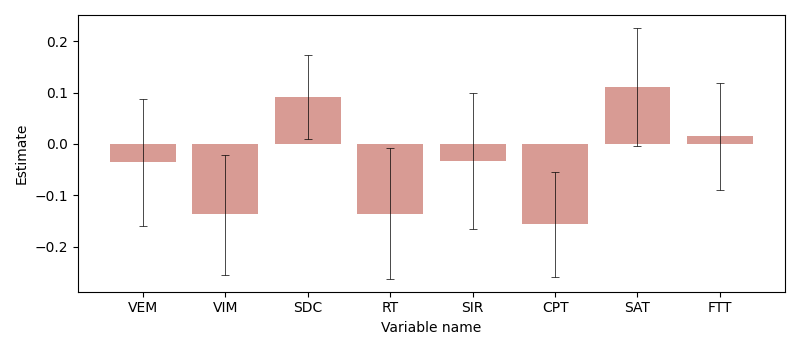

##### C: Standard deviation of the likelihood (sigma)

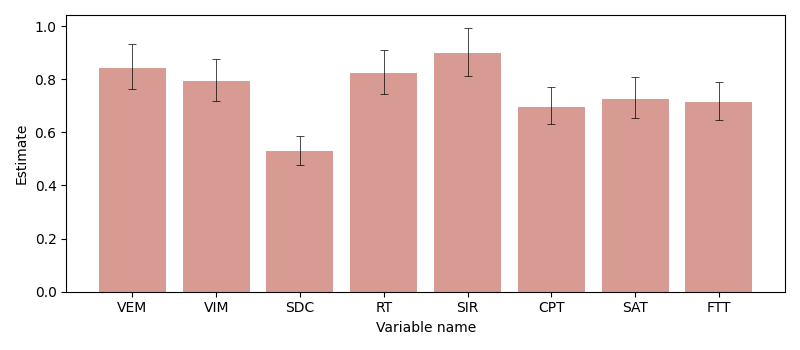

*Caption: Estimated coefficients, intercepts, and standard deviation of the likelihood (mean, the pink bars) including their credibility interval (CI: 95%) for the best-performing model (model 2c). VEM: Verbal memory recognition, VIM: Visual memory recognition, SDC: Symbol digit coding task, RT: Simple reaction time, SI: Stroop interference ratio, CPT: Continuous performance task, SAT: Shifting attention task, FTT: Finger tapping task. These same results are presented as a table in the online supplements.*

### Appendix 10: Total amount of uncertainty per outcome measure

| Test measure | Median | Lower bound (5%) | Upper bound (95%) |
| --- | --- | --- | --- |
| Verbal memory recognition | 0.875 | 0.843 | 0.927 |
| Visual memory recognition | 0.828 | 0.788 | 0.877 |
| Symbol digit coding task | 0.550 | 0.540 | 0.607 |
| Simple reaction time | 0.868 | 0.784 | 0.934 |
| Stroop interference ratio | 0.938 | 0.904 | 0.976 |
| Continuous performance task | 0.722 | 0.697 | 0.752 |
| Shifting attention task | 0.770 | 0.722 | 0.849 |
| Finger tapping task | 0.747 | 0.658 | 0.814 |

*Caption: The amount of uncertainty in individual predictions resulting from the 10-fold cross-validation procedure.*

### Appendix 11: Examples of predictions with uncertainty

To illustrate how predictions including uncertainty estimates can be implemented in clinical practice, and how uncertainty estimates can differ between patients, three of out-of-sample predictions from the best-performing model were presented for each cognitive test. These predictions were selected to represent the 5^th^ (left), 50^th^ (middle), and 95^th^ (right) percentiles of the standard deviation in the posterior predictive distribution, corresponding to certain, somewhat certain, and uncertain predictions, respectively. Visualizations show the point estimate provided by this model, the corresponding estimate of uncertainty (the posterior predictive distribution), and the true measured value, the same as in Figure 2.

Two things can be noted from these predictions. First, the uncertainty surrounding the point predictions is roughly equivalent for certain (5^th^ percentile, left) and uncertain (95^th^ percentile, right) predictions, which is in line with the high aleatoric uncertainty. Note that this aleatoric uncertainty will probably decrease in future studies, likely resulting in larger differences in the certainty of predictions for different patients. Second, the uncertainty surrounding predictions differs per outcome measure, which is in line with the differences in the variability of the outcome measures.

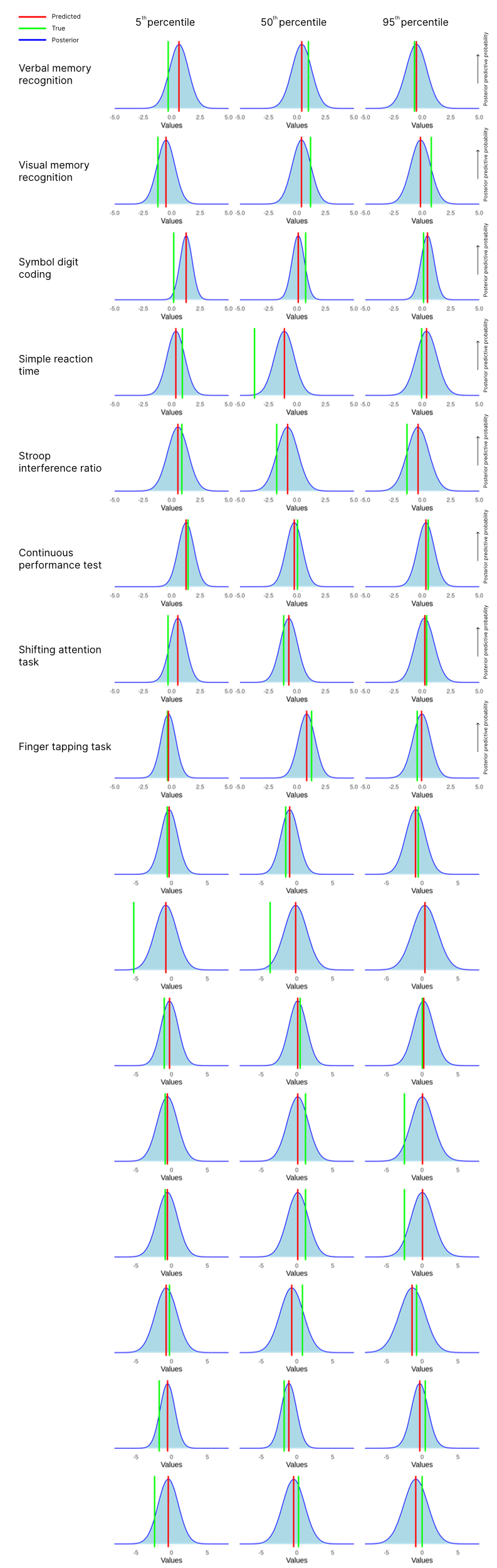

*Caption: Example predictions obtained using 10-fold cross-validation for the best-performing model (2c). Example predictions were selected to be at the 5th, 50th, and 95th percentile in terms of the amount of uncertainty in the prediction (ranging from most certain to least certain). The blue distribution represents the posterior predictive distribution resulting from the model, which represents the probability of each outcome, the red line represents the point estimate obtained from this distribution, and the green line represents the measured value.*
