## Supplementary material for "Predicting cognitive function three months after surgery in patients with a glioma": Code: Online supplement - Code.R

```
library(brms)
library("stringr")
library("readxl")
library("MLmetrics")
library("glue")
library("matrixStats")
library("writexl")
library("purrr")
library("mice")
library("cmdstanr")
library(rstan)
library("caret")
library("abind")
options(cmdstanr_write_stan_file_dir = "/home/sander/bayesianT3/")
set.seed(23)
set_cmdstan_path(path = "/home/sander/miniconda3/envs/brmst3/bin/cmdstan")
rstan_options(auto_write = TRUE)

# ulimit -s unlimited

print(cmdstan_path())

path_to_data = "/path/to/Online supplement - Example data.xlsx" #todo: Update this path
data <- read_excel(path_to_data)
colnames(data) = gsub(" ", "_", colnames(data))
  
# Settings for model fitting
do_kfold = FALSE
do_plot = FALSE
n_folds = 10 # for k-fold corss validation
n_mice = 30
library(future)
plan(multicore)
options(auto_write = TRUE)
options(mc.cores=parallel::detectCores())

# Output names
y_vars_nu = c('T3cVEM',
 'T3cVIM',
 'T3cSDC',
'T3cS1RT',
'T3cSIR32',
 'T3cCPT',
'T3cSAT',
 'T3cFTT')

y_vars = c('T3_c_VEM',
 'T3_c_VIM',
 'T3_c_SDC',
'T3_c_S1RT',
'T3_c_SIR32',
 'T3_c_CPT',
'T3_c_SAT',
 'T3_c_FTT')

predictors_to_norm = c('age_T0',
 'education_Verhage',
 'size',
 'HADS',
 'Frontal_Lobe_L',
 'Occipital_Lobe_L',
 'Parietal_Lobe_L',
 'Temporal_Lobe_L',
 'Frontal_Lobe_R',
 'Occipital_Lobe_R',
 'Parietal_Lobe_R',
 'Temporal_Lobe_R')
# Predictor names
X_vars = c('age_T0', #
 'education_Verhage', #
 'sex', #
 'lateralization_left', #
 'T0_corticosteroid', #
 'comorbidity', #
 'size', #
 'HADS', #
 'ASA_1', #
 'IDH1', #
 'WHO_high', #
 'epilepsy_or_AED', #
 'Attention_executive_memory_behavioural_SB', # may be aleviated?
 'Language_SB',
 'Paresis_motor_deficits_SB',
 'Headache_nieuw_SB',
 'Frontal_Lobe_L',
 'Occipital_Lobe_L',
 'Parietal_Lobe_L',
 'Temporal_Lobe_L',
 'Frontal_Lobe_R',
 'Occipital_Lobe_R',
 'Parietal_Lobe_R',
 'Temporal_Lobe_R',
 'glioblastoma',
 'oligodendroglioma',
'T0_c_VEM',
 'T0_c_VIM',
 'T0_c_SDC',
'T0_c_S1RT',
'T0_c_SIR32',
 'T0_c_CPT',
'T0_c_SAT',
 'T0_c_FTT')
X_vars_onlypre = c(
  'T0_c_VEM',
 'T0_c_VIM',
 'T0_c_SDC',
'T0_c_S1RT',
'T0_c_SIR32',
 'T0_c_CPT',
'T0_c_SAT',
 'T0_c_FTT'
)
# Predictor names w/o tumor type for hierarchical models
X_vars_h = c('age_T0',
 'education_Verhage',
 'sex',
 'lateralization_left',
 'T0_corticosteroid',
 'comorbidity',
 'size',
 'HADS',
 'ASA_1',
 'IDH1',
 'WHO_high',
 'epilepsy_or_AED',
 'Attention_executive_memory_behavioural_SB',
 'Language_SB',
 'Paresis_motor_deficits_SB',
 'Headache_nieuw_SB',
 'Frontal_Lobe_L',
 'Occipital_Lobe_L',
 'Parietal_Lobe_L',
 'Temporal_Lobe_L',
 'Frontal_Lobe_R',
 'Occipital_Lobe_R',
 'Parietal_Lobe_R',
 'Temporal_Lobe_R',
   'T0_c_VEM',
 'T0_c_VIM',
 'T0_c_SDC',
'T0_c_S1RT',
'T0_c_SIR32',
 'T0_c_CPT',
'T0_c_SAT',
 'T0_c_FTT'
)

# Create tumor categories
data[data$glioblastoma==TRUE, 'tumor_cat']=1
data[data$oligodendroglioma==TRUE, 'tumor_cat']=2
data[data$oligodendroglioma==FALSE & data$glioblastoma==FALSE, 'tumor_cat']=3


data['tumor_cat'] = as.factor(data$tumor_cat) #only used in hirachrical models

data[X_vars] <- lapply(data[X_vars], function(x) as.numeric(scale(x)))

data['age_times_size'] <- data['age_T0'] * data['size']
data['education_times_size'] <- data['education_Verhage'] * data['size']
data[c("age_times_size", "education_times_size")] <- lapply(data[c("age_times_size", "education_times_size")], function(x) as.numeric(scale(x)))


data_comp = data[complete.cases(data[c(y_vars)]),]
n <- nrow(data)
n_comp <- nrow(data_comp)
p <- length(y_vars)  # number of output features
# Generate balanced folds using caret
folds <- createFolds(1:n, k = n_folds, list = TRUE)
folds_comp <- createFolds(1:n_comp, k = n_folds, list = TRUE)


sum(complete.cases(data[c(X_vars,y_vars)]))
sum(complete.cases(data[c(y_vars)]))
sum(complete.cases(data[c(X_vars)]))
A = data[c(X_vars)]

# Fits the model on all data and saves the model
do_fit <- function(formula, priors, name, X_vars, y_vars_loc, complete_only, threads = 8, pp=TRUE) {
  print('Doing fit for')
  print(name)
 #formula, priors, name, X_vars, y_vars, complete_only
  if (complete_only){
      data_loc = data_comp
  }
  else {
    data_loc = data
  }
  data_loc = data_loc[c(X_vars,y_vars_loc)]

  predictorMatrix <- matrix(0, ncol(data_loc), ncol(data_loc))
  predictorMatrix[match(c(X_vars), names(data_loc)), match(c(X_vars), names(data_loc))] <- 1
  init = mice(data_loc, maxit=0)
  method = init$method
  method[!names(method) %in% c(X_vars, 'tumor_cat')] <- ''

  imp_result = mice(data_loc, predictorMatrix=predictorMatrix, method=method, m = n_mice, print = TRUE)

  data_imputed = complete(imp_result, action='long')
  data_imputed[X_vars] = lapply(data_imputed[X_vars],as.numeric)
  preproc <- preProcess(data_imputed[c(X_vars)], method = c("center", "scale"))
  data_imputed[c(X_vars)] <- predict(preproc, data_imputed[c(X_vars)])
  data_imputed = split(data_imputed, data_imputed$.imp)

  backend = "cmdstanr"
  backend = "rstan"

  fit =
    brm_multiple(
      formula = formula,
      data = data_imputed,
      prior = priors,
      control = list(adapt_delta =.99),
      family = gaussian(),
      threads = threads,
      backend = backend,
      file = name,
      file_refit = 'always',
      save_model = name,
    )
  print('Done fit')
  print(prior_summary(fit))

  fit_prior = NULL
  if (pp==TRUE){
    fit_prior =
      brm_multiple(
        formula = formula,
        data = data_imputed,
        prior = priors,
        control = list(adapt_delta =.99),
        family = gaussian(),
        threads = threads,
        backend = backend,
        file = paste0('prior', name),
        file_refit = 'always',
        # save_model = 'fit_mv',
        sample_prior = "only"
      )
    }
  print('Done prior')

  print(fit)
  if (is.list(y_vars_loc)){
    for (one_y_var in y_vars_loc){
        one_y_var = gsub("_", "", one_y_var)
        loo_res_one = LOO(fit, newdata=na.omit(data_imputed[[1]]), resp = one_y_var)
        saveRDS(loo_res_one, file = paste('loo_one_', name, one_y_var, '.rds'))
    }
  }

  loo_res = LOO(fit, newdata=na.omit(data_imputed[[1]]))
  saveRDS(loo_res, file = paste('loo_', name, '.rds'))

  return(list(fit=fit, fitprior = fit_prior))
}
# Plot prior and posterior predictive checks
plot_distributions <- function(thefit, thefitprior, thefitname){
    if (do_plot) {
      n_params <- 20
      all_variables = names(thefit$fit@sim[[1]][[1]])
      total_params <- length(all_variables)
      total_plots <- ceiling(total_params / n_params)
      for (i in 1:total_plots) {
        print(i)
        start_idx <- (i - 1) * n_params + 1
        end_idx <- min(i * n_params, total_params)
        print(start_idx)
        print(end_idx)
        variables = all_variables[start_idx:end_idx]
        toplot = plot(thefit, N=length(variables) , variable = variables,
             combo = c("trace", "dens"), ask = FALSE, plot=FALSE)
        ggsave(paste0(thefitname, i, ".jpeg"), plot=toplot[[1]], width = 20, height = 35, bg = "white")
      }
      for (variable in y_vars_nu) {
        toplot = pp_check(thefitprior, resp = variable, ndraws=30)
        ggsave(paste0(thefitname, "_prior_", variable, ".jpeg"), plot=toplot, width = 15, height = 10, bg = "white")
        toplot = pp_check(thefit, resp = variable, ndraws=30)
        ggsave(paste0(thefitname, "_posterior_", variable, ".jpeg"), plot=toplot, width = 15, height = 10, bg = "white")
      }
   }
}
# Fits the model using cross-validation
run_kfold <- function(formula, priors, name, X_vars, y_vars, complete_only, threads=64){
  # formula, priors, name, X_vars, to_predict, complete_only
  if (do_kfold) {

    if (complete_only){
        n_loc = n_comp
        folds_loc = folds_comp
        data_loc = data_comp
    }
    else {
          n_loc = n
          folds_loc = folds
          data_loc = data
    }
    data_loc = data_loc[c(X_vars,y_vars)]

    backend = "cmdstanr"
    backend = "rstan"

    # Initialize variables to store results
    all_predsmv <- matrix(NA, n_loc, length(y_vars))  # to store all predictions

    for(i in 1:n_folds) {
      print('fold')
      print(i)
      test_idx <- folds_loc[[i]]
      train_idx <- setdiff(1:n, test_idx)

      train_data <- data_loc[train_idx, , drop = FALSE]
      test_data <- data_loc[test_idx, , drop = FALSE]

      predictorMatrix <- matrix(0, ncol(data_loc), ncol(data_loc))
      predictorMatrix[match(c(X_vars), names(data_loc)), match(c(X_vars), names(data_loc))] <- 1
      init = mice(data_loc, maxit=0)
      method = init$method
      method[!names(method) %in% c(X_vars, 'tumor_cat')] <- ''

      imp_train <- mice(train_data, predictorMatrix=predictorMatrix, method=method, m = n_mice, print = TRUE)
      imp_test <- mice.mids(imp_train, newdata = test_data)

      compl_train = complete(imp_train, action='long')
      compl_train[X_vars] = lapply(compl_train[X_vars],as.numeric)
      preproc <- preProcess(train_data[c(X_vars)], method = c("center", "scale"))
      compl_train[c(X_vars)] <- predict(preproc, compl_train[c(X_vars)])
      compl_train = split(compl_train, compl_train$.imp)
      compl_test = complete(imp_test, action='long')
      compl_test[X_vars] = lapply(compl_test[X_vars],as.numeric)
      compl_test[c(X_vars)] <- predict(preproc, compl_test[c(X_vars)])
      compl_test = split(compl_test, compl_test$.imp)

      # Fit the brms model on the training data
      fitted_model =
        brm_multiple(
          formula = formula,
          data = compl_train,
          prior = priors,
          control = list(adapt_delta =.99),
          family = gaussian(),
          #future = TRUE,
          threads = threads,
          backend = backend,
          file = paste0(name, 'kfold', i),
          file_refit = 'always',
          # save_model = paste0('fit_mv', i),
          silent = 0
        )

      # Generate predictions using the fitted brms model. Average predictions of the different imputed datasets
      for (j_mice in 1:n_mice){
        comp_test_loc = compl_test[[j_mice]]
        if (j_mice == 1){
          y_pred <- posterior_predict(fitted_model, newdata = comp_test_loc)
        }
        else {
          y_pred = abind(y_pred, posterior_predict(fitted_model, newdata = comp_test_loc), along = 1)
        }
      }

      print("Saving draws")
      print(dim(y_pred))
      savedraws = list(test_idx, y_pred)
      saveRDS(savedraws, file = paste('savedraws', name, '_', i, '.rds'))
      y_pred <- colMeans(y_pred, na.rm=TRUE) # Averaging over all posterior samples resulting from the different datasets
      all_predsmv[test_idx, ] <- y_pred

    }
    # saveRDS(all_draws, file = paste('All_drawsmv', name, '.rds'))

    if (complete_only == FALSE){
      R2_mv <- sapply(1:length(y_vars), function(i) {
        print('R2 normal')
        col1 <- all_predsmv[, i, drop=FALSE]
        col2 <- data_loc[[y_vars[i]]]
        print(dim(data_loc))
        print(length(data_loc))
        valid_rows <- complete.cases(col1, col2) # Only y var of intrest needs to be complete
        correlation <- cor(col1[valid_rows], col2[valid_rows])
        correlation^2
      })
      print(R2_mv)
      saveRDS(R2_mv, file = paste('R2_', name, '.rds'))
    }

    R2_mv_comp <- sapply(1:length(y_vars), function(i) {
      print('R2 complete')
      col1 <- all_predsmv[, i, drop=FALSE]
      col2 <- data_loc[[y_vars[i]]]
      print(dim(data_loc))
      print(length(data_loc))
      valid_rows <- complete.cases(col1, data_loc[, y_vars]) # All y_vars need to be complete
      correlation <- cor(col1[valid_rows], col2[valid_rows])
      correlation^2
    })
    print(R2_mv_comp)
    saveRDS(R2_mv_comp, file = paste('R2_comp_', name, '.rds'))

  }
}

if (FALSE){
  # Server 1
  print('------------------------------------------------')
  print('New model')
  print('Multivariate')
  if (FALSE) {

    predictors = paste(X_vars, collapse ='+')
    output_vars = paste(y_vars, collapse = ', ')
    formula = bf(glue("mvbind({output_vars}) | mi() ~ {predictors}"))

    priors = c(
          reduce(map(y_vars_nu, ~ set_prior("horseshoe(3, par_ratio  = 0.5, scale_global = 0.5)", class="b", resp =.x)), `+`) ,
          reduce(map(y_vars_nu, ~ set_prior("normal(0, 3)", class = "sigma", resp = .x)),`+`) ,
          reduce(map(y_vars_nu, ~ set_prior("normal(0, 2)", class = "Intercept", resp = .x)),`+`) , # Average = -.70
          prior(lkj(6), class = rescor)
    )

    name = 't3multivariate'
    rescor = TRUE
    complete_only = FALSE
    formula = formula + set_rescor(rescor)

    fits = do_fit(formula, priors, name, X_vars, y_vars, complete_only)
    fit = fits$fit
    fitprior = fits$fitprior
    plot_distributions(fit, fitprior, name)
    run_kfold(formula, priors, name, X_vars, y_vars, complete_only)
  }
  print('------------------------------------------------')
  print('New model')
  print('Multivariate age_T0*size')
  if (TRUE) {

    X_vars_loc = c(X_vars, 'age_times_size')
    predictors = paste(X_vars_loc, collapse ='+')
    output_vars = paste(y_vars, collapse = ', ')
    formula = bf(glue("mvbind({output_vars}) | mi() ~ {predictors}"))

    priors = c(
          reduce(map(y_vars_nu, ~ set_prior("horseshoe(3, par_ratio  = 0.5, scale_global = 0.5)", class="b", resp =.x)), `+`) ,
          reduce(map(y_vars_nu, ~ set_prior("normal(0, 3)", class = "sigma", resp = .x)),`+`) ,
          reduce(map(y_vars_nu, ~ set_prior("normal(0, 2)", class = "Intercept", resp = .x)),`+`) , # Average = -.70
          prior(lkj(6), class = rescor)
    )

    name = 't3multivariate age times size'
    rescor = TRUE
    complete_only = FALSE
    formula = formula + set_rescor(rescor)

    # fits = do_fit(formula, priors, name, X_vars_loc, y_vars, complete_only)
    # fit = fits$fit
    # fitprior = fits$fitprior
    # plot_distributions(fit, fitprior, name)
    run_kfold(formula, priors, name, X_vars_loc, y_vars, complete_only)
  }
  print('------------------------------------------------')
  print('New model')
  print('Multivariate education_Verhage*size')
  if (FALSE) {

    X_vars_loc = c(X_vars, 'education_times_size')
    predictors = paste(X_vars_loc, collapse ='+')
    output_vars = paste(y_vars, collapse = ', ')
    formula = bf(glue("mvbind({output_vars}) | mi() ~ {predictors}"))

    priors = c(
          reduce(map(y_vars_nu, ~ set_prior("horseshoe(3, par_ratio  = 0.5, scale_global = 0.5)", class="b", resp =.x)), `+`) ,
          reduce(map(y_vars_nu, ~ set_prior("normal(0, 3)", class = "sigma", resp = .x)),`+`) ,
          reduce(map(y_vars_nu, ~ set_prior("normal(0, 2)", class = "Intercept", resp = .x)),`+`) , # Average = -.70
          prior(lkj(6), class = rescor)
    )

    name = 't3multivariate education times size2'
    rescor = TRUE
    complete_only = FALSE
    formula = formula + set_rescor(rescor)

    fits = do_fit(formula, priors, name, X_vars_loc, y_vars, complete_only)
    fit = fits$fit
    fitprior = fits$fitprior
    plot_distributions(fit, fitprior, name)
    run_kfold(formula, priors, name, X_vars_loc, y_vars, complete_only)
  }
}

if (FALSE) {
  print('------------------------------------------------')
  print('New model')
  print('No pooling ') # this one
  if (FALSE) {

    predictors = paste(X_vars_h, collapse ='+')
    output_vars = paste(y_vars, collapse = ', ')
    formula = bf(glue("mvbind({output_vars}) | mi() ~ ({predictors}) * as.factor(tumor_cat)"))

    priors = c(
      reduce(map(y_vars_nu, ~ set_prior("horseshoe(3, par_ratio  = 0.5, scale_global = 0.5)", class="b", resp =.x)), `+`) ,
      reduce(map(y_vars_nu,~ set_prior("normal(0, 3)", class = "sigma", resp = .x)),`+`) ,
      reduce(map(y_vars_nu,~ set_prior("normal(0, 2)", class = "Intercept", resp = .x)),`+`),
      prior(lkj(6), class = rescor))

    name = 't3No pooling'
    rescor = TRUE
    complete_only = FALSE
    formula = formula + set_rescor(rescor)

    fits = do_fit(formula, priors, name, c(X_vars_h, 'tumor_cat'), y_vars, complete_only)
    fit = fits$fit
    fitprior = fits$fitprior
    plot_distributions(fit, fitprior, name)
    run_kfold(formula, priors, name, c(X_vars_h, 'tumor_cat'), y_vars, complete_only)
  }
  print('------------------------------------------------')
  print('New model')
  print('No pooling age_T0*size') # this one
  if (FALSE) {

    X_vars_loc = c(X_vars_h, 'age_times_size')
    predictors = paste(X_vars_loc, collapse ='+')
    output_vars = paste(y_vars, collapse = ', ')
    formula = bf(glue("mvbind({output_vars}) | mi() ~ ({predictors}) * as.factor(tumor_cat)"))

    priors = c(
      reduce(map(y_vars_nu, ~ set_prior("horseshoe(3, par_ratio  = 0.5, scale_global = 0.5)", class="b", resp =.x)), `+`) ,
      reduce(map(y_vars_nu,~ set_prior("normal(0, 3)", class = "sigma", resp = .x)),`+`) ,
      reduce(map(y_vars_nu,~ set_prior("normal(0, 2)", class = "Intercept", resp = .x)),`+`),
      prior(lkj(6), class = rescor))

    name = 't3No pooling age_times_size'
    rescor = TRUE
    complete_only = FALSE
    formula = formula + set_rescor(rescor)

    fits = do_fit(formula, priors, name, c(X_vars_loc, 'tumor_cat'), y_vars, complete_only)
    fit = fits$fit
    fitprior = fits$fitprior
    plot_distributions(fit, fitprior, name)
    run_kfold(formula, priors, name, c(X_vars_loc, 'tumor_cat'), y_vars, complete_only)
  }
  print('------------------------------------------------')
  print('New model')
  print('No pooling education_Verhage*size') # this one
  if (FALSE) {

    X_vars_loc = c(X_vars_h, 'education_times_size')
    predictors = paste(X_vars_loc, collapse ='+')
    output_vars = paste(y_vars, collapse = ', ')
    formula = bf(glue("mvbind({output_vars}) | mi() ~ ({predictors}) * as.factor(tumor_cat)"))

    priors = c(
      reduce(map(y_vars_nu, ~ set_prior("horseshoe(3, par_ratio  = 0.5, scale_global = 0.5)", class="b", resp =.x)), `+`) ,
      reduce(map(y_vars_nu,~ set_prior("normal(0, 3)", class = "sigma", resp = .x)),`+`) ,
      reduce(map(y_vars_nu,~ set_prior("normal(0, 2)", class = "Intercept", resp = .x)),`+`),
      prior(lkj(6), class = rescor))

    name = 't3No pooling education_times_size'
    rescor = TRUE
    complete_only = FALSE
    formula = formula + set_rescor(rescor)

    # fits = do_fit(formula, priors, name, c(X_vars_loc, 'tumor_cat'), y_vars, complete_only)
    # fit = fits$fit
    # fitprior = fits$fitprior
    # plot_distributions(fit, fitprior, name)
    run_kfold(formula, priors, name, c(X_vars_loc, 'tumor_cat'), y_vars, complete_only)
  }
}

# todo: always run this one seperately or at the end as threading does not work
# Seperate script
if (FALSE){
  print('------------------------------------------------')
  print('New model')
  print('Partial pooling') #1
  if (FALSE) {

    # Create formula
    predictors = paste(X_vars_h, collapse ='+')
    output_vars = paste(y_vars, collapse = ', ')
    formula = bf(glue("mvbind({output_vars}) | mi() ~ {predictors}  + (1 + {predictors} || tumor_cat)"))

    priors = c(
      reduce(map(y_vars_nu, ~ set_prior("horseshoe(3, par_ratio  = 0.5, scale_global = 0.5)", class="b", resp =.x)), `+`) ,
      reduce(map(y_vars_nu,~ set_prior("normal(0, 3)", class = "sigma", resp = .x)),`+`) ,
      reduce(map(y_vars_nu,~ set_prior("normal(0, 2)", class = "Intercept", resp = .x)),`+`),
      reduce(map(y_vars_nu,~ set_prior("normal(0, 0.5)", class = "sd", resp = .x)),`+`) ,
      prior(lkj(6), class = rescor),
      prior(lkj(6), class = cor, group=tumor_cat)
    )

    name = 't3Partial pooling2 0_5'
    rescor = TRUE
    complete_only = FALSE
    formula = formula + set_rescor(rescor)

    fits = do_fit(formula, priors, name, c(X_vars_h, 'tumor_cat'), y_vars, complete_only, threads=NULL)
    # fits = do_fit(formula, priors, name, c(X_vars_h, 'tumor_cat'), y_vars, complete_only)
    fit = fits$fit
    fitprior = fits$fitprior
    #plot_distributions(fit, fitprior, name)
    #run_kfold(formula, priors, name, c(X_vars_h, 'tumor_cat'), y_vars, complete_only, NULL)
  }
  print('------------------------------------------------')
  print('New model')
  print('Partial pooling age_T0*size') #1
  if (FALSE) {

    # Create formula
    X_vars_loc = c(X_vars_h, 'age_times_size')
    predictors = paste(X_vars_loc, collapse ='+')
    output_vars = paste(y_vars, collapse = ', ')
    formula = bf(glue("mvbind({output_vars}) | mi() ~ {predictors}  + (1 + {predictors} || tumor_cat)"))
    # Instead of | you may use || in grouping terms to prevent correlations from being modeled.

    priors = c(
      reduce(map(y_vars_nu, ~ set_prior("horseshoe(3, par_ratio  = 0.5, scale_global = 0.5)", class="b", resp =.x)), `+`) ,
      reduce(map(y_vars_nu,~ set_prior("normal(0, 3)", class = "sigma", resp = .x)),`+`) ,
      reduce(map(y_vars_nu,~ set_prior("normal(0, 2)", class = "Intercept", resp = .x)),`+`),
      reduce(map(y_vars_nu,~ set_prior("normal(0, 0.5)", class = "sd", resp = .x)),`+`) ,
      prior(lkj(6), class = rescor),
      prior(lkj(6), class = cor, group=tumor_cat)
    )

    name = 't3Partial pooling2 age_times_size 0_5'
    rescor = TRUE
    complete_only = FALSE
    formula = formula + set_rescor(rescor)

    fits = do_fit(formula, priors, name, c(X_vars_loc, 'tumor_cat'), y_vars, complete_only, threads=NULL)
    # fits = do_fit(formula, priors, name, c(X_vars_h, 'tumor_cat'), y_vars, complete_only)
    fit = fits$fit
    fitprior = fits$fitprior
    #plot_distributions(fit, fitprior, name)
    #run_kfold(formula, priors, name, c(X_vars_loc, 'tumor_cat'), y_vars, complete_only, NULL)
  }
  print('------------------------------------------------')
  print('New model')
  print('Partial pooling education_Verhage*size') #1
  if (FALSE) {

    # Create formula
    X_vars_loc = c(X_vars_h, 'education_times_size')
    predictors = paste(X_vars_loc, collapse ='+')
    output_vars = paste(y_vars, collapse = ', ')
    formula = bf(glue("mvbind({output_vars}) | mi() ~ {predictors}  + (1 + {predictors} || tumor_cat)"))
    # Instead of | you may use || in grouping terms to prevent correlations from being modeled.

    priors = c(
      reduce(map(y_vars_nu, ~ set_prior("horseshoe(3, par_ratio  = 0.5, scale_global = 0.5)", class="b", resp =.x)), `+`) ,
      reduce(map(y_vars_nu,~ set_prior("normal(0, 3)", class = "sigma", resp = .x)),`+`) ,
      reduce(map(y_vars_nu,~ set_prior("normal(0, 2)", class = "Intercept", resp = .x)),`+`),
      reduce(map(y_vars_nu,~ set_prior("normal(0, 0.5)", class = "sd", resp = .x)),`+`) ,
      prior(lkj(6), class = rescor),
      prior(lkj(6), class = cor, group=tumor_cat)
    )


    name = 't3Partial pooling2 education_times_size 0_5'
    rescor = TRUE
    complete_only = FALSE
    formula = formula + set_rescor(rescor)

    fits = do_fit(formula, priors, name, c(X_vars_loc, 'tumor_cat'), y_vars, complete_only, threads=NULL)
    # fits = do_fit(formula, priors, name, c(X_vars_h, 'tumor_cat'), y_vars, complete_only)
    fit = fits$fit
    fitprior = fits$fitprior
    #plot_distributions(fit, fitprior, name)
    #run_kfold(formula, priors, name, c(X_vars_loc, 'tumor_cat'), y_vars, complete_only, NULL)
  }

}


print('------------------------------------------------')
print('Extra comparisons')
print('------------------------------------------------')
print('------------------------------------------------')
print('------------------------------------------------')
print('------------------------------------------------')
if (FALSE) {
  # Server 1
  print('------------------------------------------------')
  print('New model')
  print('Multivariate onlypreop')
  if (TRUE) {

    predictors = paste(X_vars_onlypre, collapse ='+')
    output_vars = paste(y_vars, collapse = ', ')
    formula = bf(glue("mvbind({output_vars}) | mi() ~ {predictors}"))

    priors = c(
          reduce(map(y_vars_nu, ~ set_prior("horseshoe(3, par_ratio  = 0.5, scale_global = 0.5)", class="b", resp =.x)), `+`) ,
          reduce(map(y_vars_nu, ~ set_prior("normal(0, 3)", class = "sigma", resp = .x)),`+`) ,
          reduce(map(y_vars_nu, ~ set_prior("normal(0, 2)", class = "Intercept", resp = .x)),`+`) , # Average = -.70
          prior(lkj(6), class = rescor)
    )

    name = 't3multivariate onlypreop2'
    rescor = TRUE
    complete_only = FALSE
    formula = formula + set_rescor(rescor)

    fits = do_fit(formula, priors, name, X_vars, y_vars, complete_only)
    fit = fits$fit
    fitprior = fits$fitprior
    plot_distributions(fit, fitprior, name)
    run_kfold(formula, priors, name, X_vars, y_vars, complete_only)
  }
  print('------------------------------------------------')
  print('New model')
  print('No pooling onlypreop') # this one
  if (FALSE) {

    predictors = paste(X_vars_onlypre, collapse ='+')
    output_vars = paste(y_vars, collapse = ', ')
    formula = bf(glue("mvbind({output_vars}) | mi() ~ ({predictors}) * as.factor(tumor_cat)"))

    priors = c(
      reduce(map(y_vars_nu, ~ set_prior("horseshoe(3, par_ratio  = 0.5, scale_global = 0.5)", class="b", resp =.x)), `+`) ,
      reduce(map(y_vars_nu,~ set_prior("normal(0, 3)", class = "sigma", resp = .x)),`+`) ,
      reduce(map(y_vars_nu,~ set_prior("normal(0, 2)", class = "Intercept", resp = .x)),`+`),
      prior(lkj(6), class = rescor))

    name = 't3No pooling onlypreop'
    rescor = TRUE
    complete_only = FALSE
    formula = formula + set_rescor(rescor)

    fits = do_fit(formula, priors, name, c(X_vars_h, 'tumor_cat'), y_vars, complete_only)
    fit = fits$fit
    fitprior = fits$fitprior
    plot_distributions(fit, fitprior, name)
    run_kfold(formula, priors, name, c(X_vars_h, 'tumor_cat'), y_vars, complete_only)
  }
  print('------------------------------------------------')
  print('New model')
  print('Partial pooling onlypreop') #1
  if (FALSE) {

    # Create formula
    predictors = paste(X_vars_onlypre, collapse ='+')
    output_vars = paste(y_vars, collapse = ', ')
    formula = bf(glue("mvbind({output_vars}) | mi() ~ {predictors} + (1 + {predictors} || tumor_cat)"))

    priors = c(
      reduce(map(y_vars_nu, ~ set_prior("horseshoe(3, par_ratio  = 0.5, scale_global = 0.5)", class="b", resp =.x)), `+`) ,
      reduce(map(y_vars_nu,~ set_prior("normal(0, 3)", class = "sigma", resp = .x)),`+`) ,
      reduce(map(y_vars_nu,~ set_prior("normal(0, 2)", class = "Intercept", resp = .x)),`+`),
      reduce(map(y_vars_nu,~ set_prior("normal(0, 0.5)", class = "sd", resp = .x)),`+`) ,
      prior(lkj(6), class = rescor)#,
      #prior(lkj(6), class = cor, group=tumor_cat)
    )

    name = 't3Partial pooling2 0_5 onlypreop'
    rescor = TRUE
    complete_only = FALSE
    formula = formula + set_rescor(rescor)

    # fits = do_fit(formula, priors, name, c(X_vars_h, 'tumor_cat'), y_vars, complete_only, threads=NULL)
    # fit = fits$fit
    # fitprior = fits$fitprior
    #plot_distributions(fit, fitprior, name)
    run_kfold(formula, priors, name, c(X_vars_h, 'tumor_cat'), y_vars, complete_only, NULL)
  }
}

print('------------------------------------------------')
print('Sensitivity test')
print('------------------------------------------------')
print('------------------------------------------------')
print('------------------------------------------------')
print('------------------------------------------------')
if (TRUE){
  print('------------------------------------------------')
  print('New model')
  print('No pooling education_Verhage*size default') # this one
  if (FALSE) {

    X_vars_loc = c(X_vars_h, 'education_times_size')
    predictors = paste(X_vars_loc, collapse ='+')
    output_vars = paste(y_vars, collapse = ', ')
    formula = bf(glue("mvbind({output_vars}) | mi() ~ ({predictors}) * as.factor(tumor_cat)"))

    priors = c(
      reduce(map(y_vars_nu, ~ set_prior("horseshoe(3, par_ratio  = 0.5, scale_global = 0.5)", class="b", resp =.x)), `+`) ,
      reduce(map(y_vars_nu,~ set_prior("normal(0, 3)", class = "sigma", resp = .x)),`+`) ,
      reduce(map(y_vars_nu,~ set_prior("normal(0, 2)", class = "Intercept", resp = .x)),`+`),
      prior(lkj(6), class = rescor))
    priors = c()

    name = 't3No pooling education_times_size default'
    rescor = TRUE
    complete_only = FALSE
    formula = formula + set_rescor(rescor)

    fits = do_fit(formula, priors, name, c(X_vars_loc, 'tumor_cat'), y_vars, complete_only, pp=FALSE)
    fit = fits$fit
    fitprior = fits$fitprior
    plot_distributions(fit, fitprior, name)
    # run_kfold(formula, priors, name, c(X_vars_loc, 'tumor_cat'), y_vars, complete_only)
  }
  print('------------------------------------------------')
  print('New model')
  print('No pooling education_Verhage*size no HS') # this one
  if (FALSE) {

    X_vars_loc = c(X_vars_h, 'education_times_size')
    predictors = paste(X_vars_loc, collapse ='+')
    output_vars = paste(y_vars, collapse = ', ')
    formula = bf(glue("mvbind({output_vars}) | mi() ~ ({predictors}) * as.factor(tumor_cat)"))

    priors = c(
      # reduce(map(y_vars_nu, ~ set_prior("horseshoe(3, par_ratio  = 0.5, scale_global = 0.5)", class="b", resp =.x)), `+`) ,
      reduce(map(y_vars_nu,~ set_prior("normal(0, 3)", class = "sigma", resp = .x)),`+`) ,
      reduce(map(y_vars_nu,~ set_prior("normal(0, 2)", class = "Intercept", resp = .x)),`+`),
      prior(lkj(6), class = rescor))

    name = 't3No pooling education_times_size no HS'
    rescor = TRUE
    complete_only = FALSE
    formula = formula + set_rescor(rescor)

    fits = do_fit(formula, priors, name, c(X_vars_loc, 'tumor_cat'), y_vars, complete_only, pp=FALSE)
    fit = fits$fit
    fitprior = fits$fitprior
    plot_distributions(fit, fitprior, name)
    # run_kfold(formula, priors, name, c(X_vars_loc, 'tumor_cat'), y_vars, complete_only)
  }
  print('------------------------------------------------')
  print('New model')
  print('No pooling education_Verhage*size only HS') # this one
  if (TRUE) {

    X_vars_loc = c(X_vars_h, 'education_times_size')
    predictors = paste(X_vars_loc, collapse ='+')
    output_vars = paste(y_vars, collapse = ', ')
    formula = bf(glue("mvbind({output_vars}) | mi() ~ ({predictors}) * as.factor(tumor_cat)"))

    priors = c(
      reduce(map(y_vars_nu, ~ set_prior("horseshoe(3, par_ratio  = 0.5, scale_global = 0.5)", class="b", resp =.x)), `+`) )#,
      #reduce(map(y_vars_nu,~ set_prior("normal(0, 3)", class = "sigma", resp = .x)),`+`) ,
      #reduce(map(y_vars_nu,~ set_prior("normal(0, 2)", class = "Intercept", resp = .x)),`+`),
      #prior(lkj(6), class = rescor))

    name = 't3No pooling education_times_size only HS'
    rescor = TRUE
    complete_only = FALSE
    formula = formula + set_rescor(rescor)

    fits = do_fit(formula, priors, name, c(X_vars_loc, 'tumor_cat'), y_vars, complete_only, pp=FALSE)
    fit = fits$fit
    fitprior = fits$fitprior
    plot_distributions(fit, fitprior, name)
    # run_kfold(formula, priors, name, c(X_vars_loc, 'tumor_cat'), y_vars, complete_only)
  }
}
```
